# Evolution of COVID-19 cases in selected low- and middle-income countries: have they peaked due to high levels of infection and immunity?

**DOI:** 10.1101/2020.09.26.20201814

**Authors:** Axel S. Lexmond, Carlijn J.A. Nouwen, Othmane Fourtassi, J. Paul Callan

**Author notes:** Corresponding authors., (C.J.A.N.); (J.P.C.). These authors contributed equally to this work (Axel S. Lexmond, Carlijn J.A. Nouwen, J. Paul Callan).

## Abstract

COVID-19 cases have peaked and declined rapidly in many low- and middle-income countries in recent months, in some cases after control measures were relaxed. For 10 such countries, the hypothesis that COVID-19 cases have declined mainly through low susceptibility levels, stemming largely from high levels of infection leading to (at least temporary) immunity, warrants serious consideration. The Reed-Frost model, perhaps the simplest description for the evolution of cases in an epidemic, with only a few constant parameters, fits the observed case data remarkably well, and yields parameter values that are reasonable. The model results give infection levels of 45% and 79%, above the herd immunity threshold for each country under their current social distancing conditions. Reproduction numbers range between 1.4 and 2.0, indicating that epidemic curves were “flattened” but not “suppressed”. Between 0.05% and 2.86% of cases have been detected according to the estimates – values which are consistent with findings from serological studies. Overall infection fatality ratios for two of three countries studied are lower than expected from reported infection fatality ratios by age (which are based on studies of several high-income countries). COVID-19 may have lower age-specific fatality risks in some countries, due to differences in immune-response, prior exposure to coronaviruses, disease characteristics or other factors. We find that the hypothesis of control through low susceptibility would not have fit the evolution of reported cases in several European countries, even just after the initial peaks; instead, these countries reduced COVID-19 cases initially through disease control measures – and subsequent resurgences of cases obviously prove that those countries have infection levels well below those required for herd immunity. Our hypothesis that the 10 countries we studied have low susceptibility levels should now be tested further through immunity studies, and efforts should continue to determine the duration and extent of immunity to SARS-CoV-2 after infection.

## Introduction

Figure 1, based on a similar one produced by researchers at Imperial College London *[1]*, illustrates how case numbers are expected to evolve during an epidemic under different conditions. The green curve shows expected cases for an uncontrolled outbreak. This curve has three main features: initial exponential growth in new cases, followed by a single peak as the cumulative cases reach a level at which the remaining susceptible population is not large enough to sustain further growth, and an exponential decline in new cases. The curve shape is characterised by two numbers. First, the basic reproduction number, *R*_*0*_, the average number of new infections caused by each current infected individual, at the outset of the disease outbreak when few people have been infected yet. Second, the mean generation time, *t*_*g*_, the average time between infection of one person and when that person infects other people. The peak of the curve is reached as the proportion of the population that remains susceptible drops below *1/R*_*0*_.

**Figure 1.**
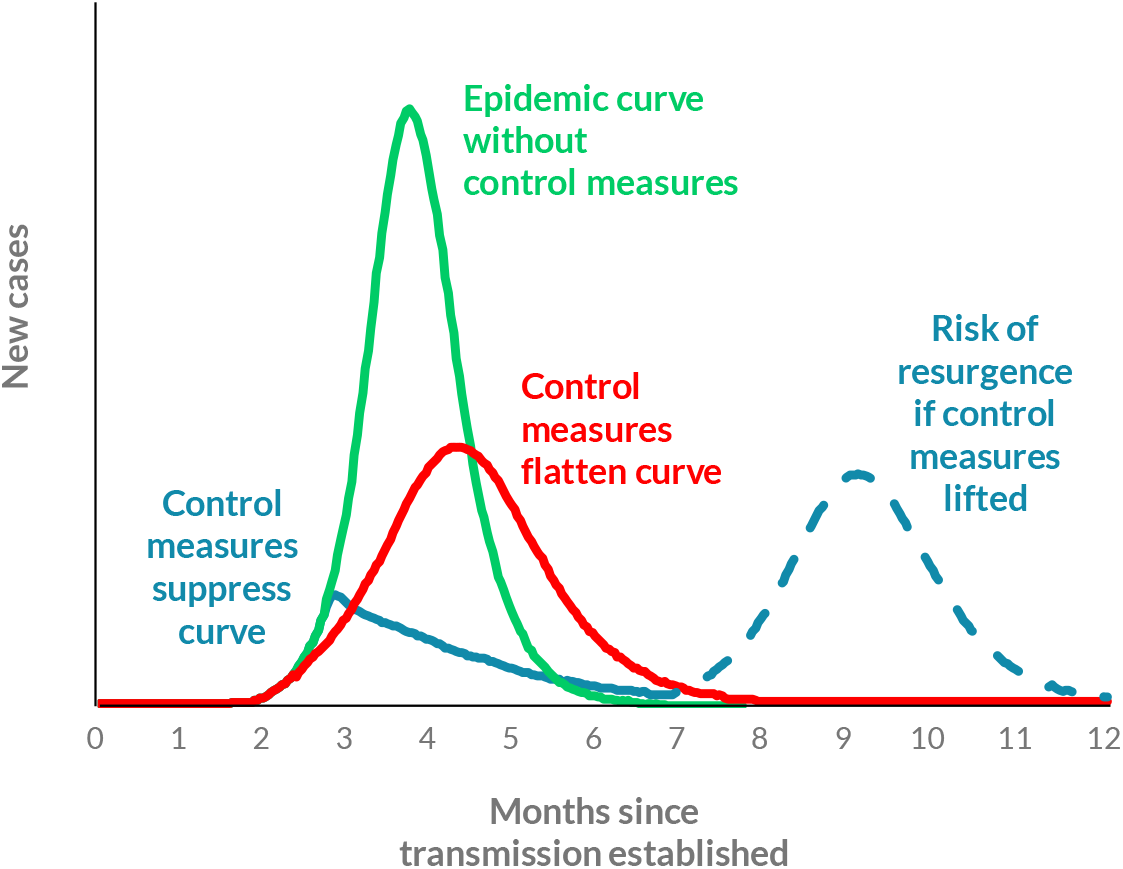
Typical disease outbreak curves in different scenarios [1].

The red curve shows expected cases when governments and people take measures to control disease spread. The red curve displays largely the same features as the green – disease spread is only halted by high infection levels – but the curve is “flattened”, with cases spread out more over time and fewer cases at the peak. In this case, disease control measures reduce the reproduction number to an effective basic reproduction number, *R*_*0_e*_ *[2]*. However, this number remains above 1, and the curve peaks when the proportion of the population that is susceptible reduces to *1/R*_*0_e*_. Note that the “effective herd immunity threshold” (which is *1–1/R*_*0_e*_) is not a fixed quantity: it depends on disease control measures and will increase when control measures are lifted; and only when the “full” or “natural” herd immunity threshold, *1–1/R*_*0*_, is reached, through infection and/or vaccination, will the disease be constrained even in the absence of control measures.

The blue curve shows the evolution of cases when disease control measures are sufficient to bring *R*_*0_e*_ below 1. The curve is “crushed” or “suppressed”; the initial exponential growth in case numbers is halted and cases decline (almost always at a slower rate than for the green or red curves). In this case, because most people are still susceptible to the disease, it is possible for the disease to return if containment measures later allow *R*_*0_e*_ to increase above 1, as shown in the dashed part of the blue curve in Figure 1.

### Patterns in reported cases

Reported cases in several low- and middle-income countries (LMICs) have evolved in a manner that is very similar to the red and green curves of Figure 1. We show the 7-day rolling average of reported new cases for 10 such countries in Figure 2.

**Figure 2.**
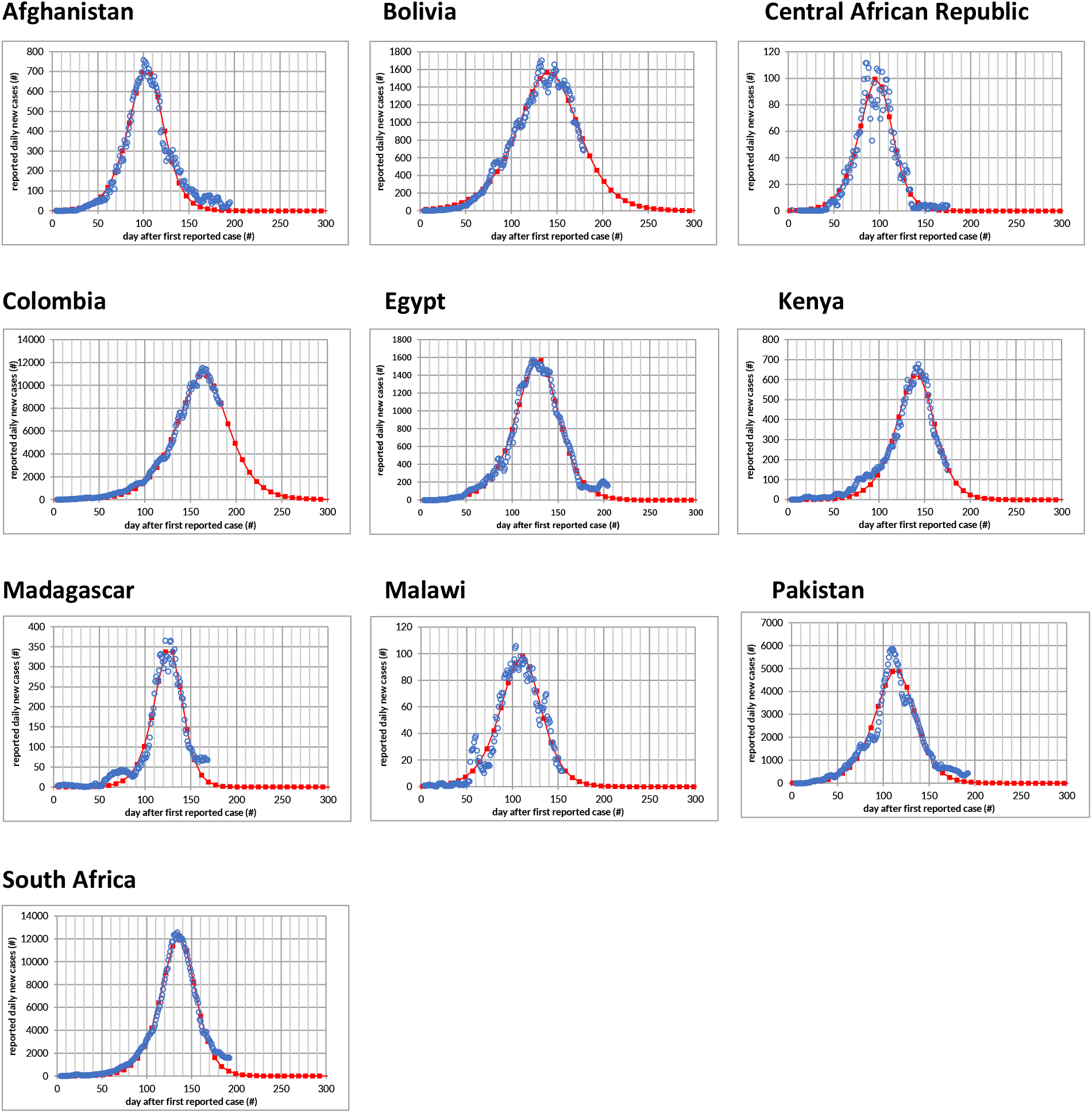
COVID-19 reported cases in 10 selected low- and middle-income countries, together with best-fit curves from simple disease spread model. Blue circles show daily reported new cases of COVID-19 (based on a 7-day rolling average). Red squares (joined by straight lines) show best-fit outputs from linearized Reed-Frost model.

Other countries show similar patterns; we have chosen to study a subset with the clearest similarities to expected outbreak curves like the red and green curves of Figure 1. In all these countries, the reported cases have (1) grown exponentially, (2) reached a single clear peak and (3) declined exponentially. Regulations were most stringent, and compliance was greatest, in most of these countries, after the designation of the global pandemic in March and have relaxed to varying degrees in recent months – but cases continued to decline. None of these countries has reported a significant increase in new cases after the peak that would indicate a second wave (although cases in some countries have only recently passed the peak). Together, these observations point to a hypothesis that the outbreaks in these countries have reached sufficiently low levels of remaining susceptibility, and that the recently observed declines in new cases are because many people are not susceptible– at least temporarily.

However, the numbers of cases in other countries – including most high-income countries (HICs) but also some LMICs – show patterns that are much different. Figure 3 shows 7-day rolling averages of reported cases for 6 such comparison countries. In these countries, cases have evolved in a manner that is similar to the first part of the blue curve of Figure 1. There have been peaks in numbers of reported cases, yet the decline is often longer and slower than the red or green curves would suggest. In some countries, there have been resurgences in cases, indicating that the initial suppression was not due to low levels of susceptibility.

**Figure 3.**
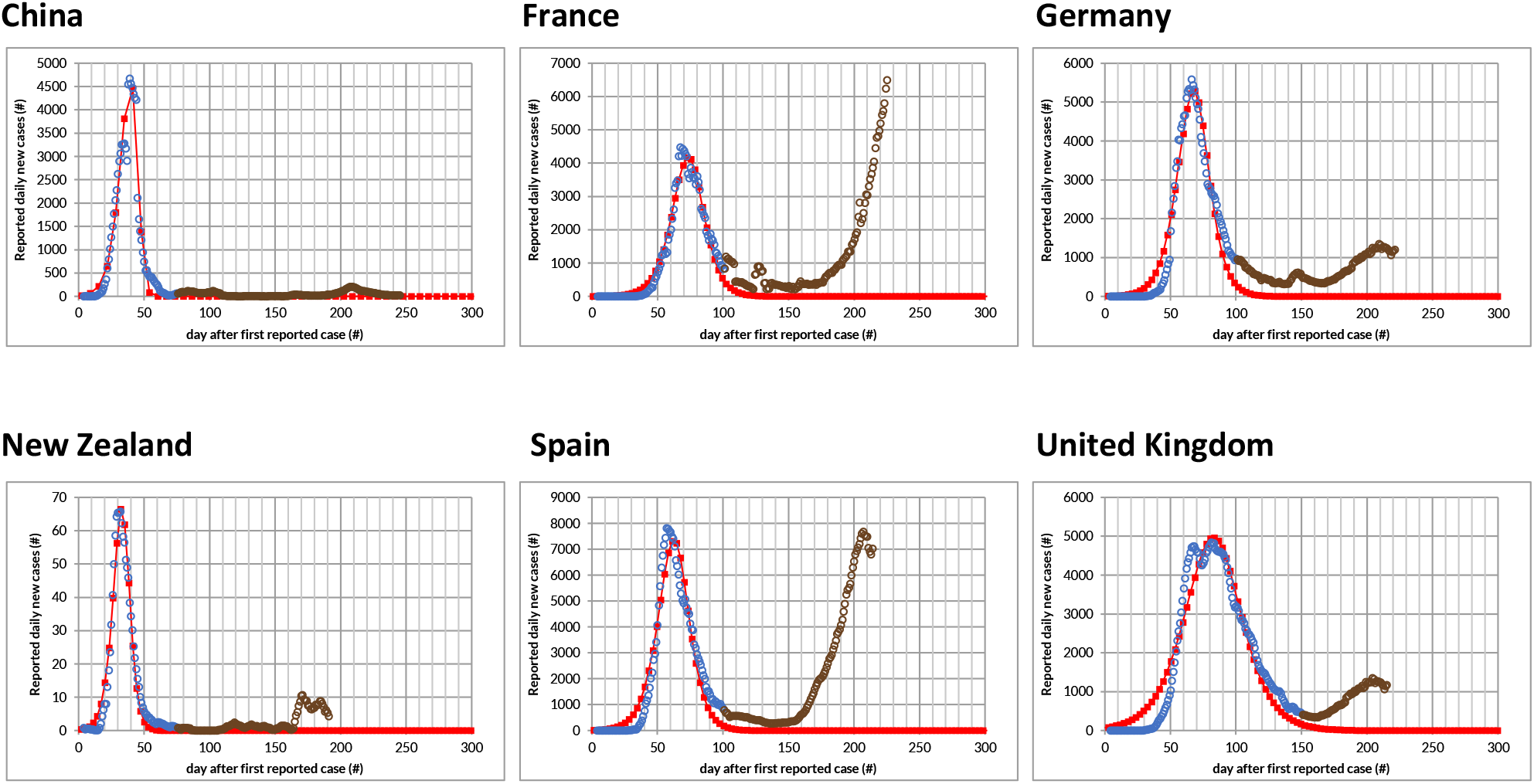
COVID-19 reported cases in 6 selected comparison countries, together with best-fit curves from test of simple disease outbreak model. Blue and brown circles show daily reported new cases of COVID-19 (based on a 7-day rolling average). Red squares (joined by straight lines) show best-fit outputs from linearized Reed-Frost model, fit only to the blue circles in the first peaks in cases.

### Fit with disease outbreak model and estimation of outbreak parameters

We test our hypothesis that the disease dynamics in these 10 countries has been driven primarily by susceptibility levels, by using a simple disease outbreak model and fitting to the reported cases. The outbreak model is a linearised Reed-Frost model *[3]*, the textbook deterministic mathematical model for an epidemic. The curves produced by this model depend on just two parameters, namely the effective basic reproduction number (*R*_*0_e*_) and the mean generation time (*t*_*g*_). Expected reported cases are calculated by scaling the Reed-Frost model’s results by a detection rate (*p*) *[4]*. The parameters that produce the best-fit curve for reported cases in each country are determined partly analytically from the observed data and partly from least squares regression. Our model parameters (*R*_*0_e*_, *t*_*g*_ and *p*) are constant over time – a beneficial assumption in that it avoids having too many free parameters, which might lead to good fits even if the model incorrectly describes the disease dynamics. Furthermore, in most of the countries studied, reported cases and reported deaths have followed similar trends (with changes in deaths lagging the corresponding changes in cases), even for countries with very low absolute numbers of reported cases and deaths – which suggests that the shapes of the curves likely reflect trends in actual cases and deaths, and that detection rates for both do not vary wildly over time *[5]*.

The best-fit curves are shown with red squares in Figure 2, for each of the 10 LMICs studied, and the corresponding parameters are presented in Table 1. For South Africa, *R*_*0_e*_ and *t*_*g*_ are calculated from the slopes and width of the observed data, and *p* is calculated from the sum of reported cases up to the peak divided by the total population times the expected proportion of population infected at the peak. For the other 9 countries, we use the value of *t*_*g*_ calculated for South Africa, and determine *R*_*0_e*_ and *p* from fitting to the observed case data. The fits are close, with R-squared goodness-of-fit measures between 0.94 and 1.00. (The fits are somewhat less good for Central African Republic and Malawi, due to very low numbers of reported cases, and for Pakistan, due to undulations in reported cases which may signal variations in *R*_*0_e*_ or *p* which the model assumes are constant.) These results demonstrate that the observed case patterns can indeed very accurately be described by an exponential outbreak halted by declining numbers of people still susceptible to infection.

**Table 1.**
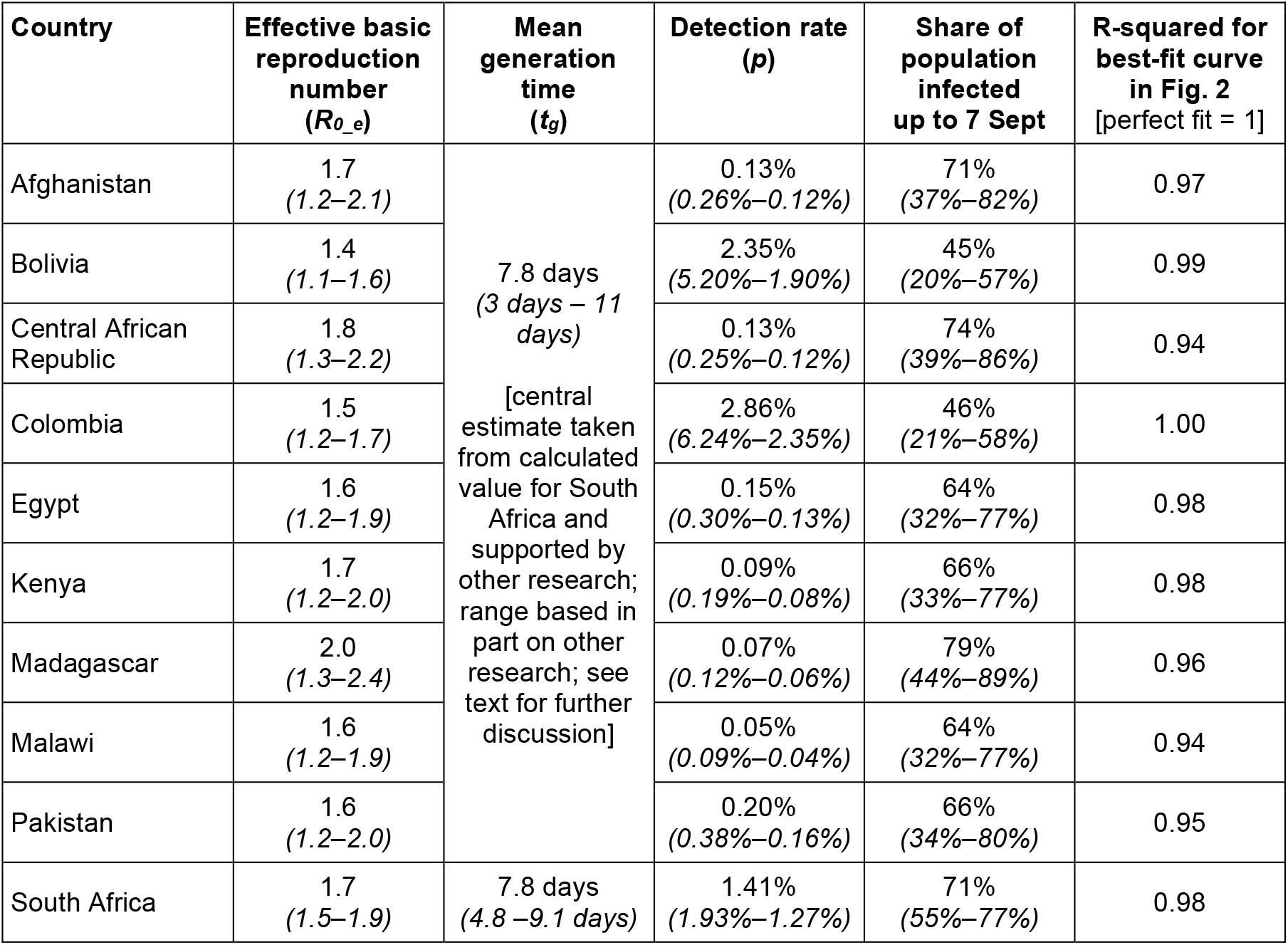
Outbreak parameters for selected countries – effective basic reproduction number (R_0_e_), mean generation time (t_g_) and case detection rate (p) – and implied share of population infected as of 7 September. The best estimates for the parameters correspond to the best-fit curves shown in Figure 2, and ranges of possible values for each parameter are given in parentheses (and as discussed in the text, are generated by letting *t*_*g*_ vary from 3 days to 11 days). The share of population infected up to 7 September is determined from the model outputs. The table also shows the values of the goodness-of-fit measure R-squared for the best-fit curves in Figure 2.

Table 1 presents best estimates for *R*_*0_e*_, *p* and the infection level (on 7 September), together with ranges of possible values (in parentheses). There are fairly wide ranges of reasonably possible values because the effects of *R*_*0_e*_ and *t*_*g*_ on the observed case curves are hard to distinguish – especially when the observed data has more “noise” or when the observed data does not include many points after the peak, and when values of *R*_*0_e*_ are close to 1 *[6]*. For South Africa, *R*_*0_e*_ has a best-estimate value of 1.74 (and a range of possible values between 1.45 and 1.90); the corresponding value of *t*_*g*_ is 7.8 days (4.8 days – 9.1 days) and of *p* is 1.41% (1.93% – 1.27%). For the other 9 countries, we determine best estimates of *R*_*0_e*_ and *p* by assuming that t_g_ = 7.8 days and fitting model outputs to data on reported cases. The value of *t*_*g*_ = 7.8 days is consistent with studies of serial intervals – the time between illness onset in successive cases in a transmission chain, whose mean value should equal the mean generation time *t*_*g*_ – by Ali *et al*. and others *[7]*, and close to the values used in other COVID-19 models *[8]*. We also conduct sensitivity analyses, determining ranges of possible values of *R*_*0_e*_ and *p* if *t*_*g*_ varies between 3 days and 11 days (wider than the range of possible values suggested in the literature *[7,8]*), and finding the values of *R*_*0_e*_ and *p* that produce the best fits to the reported cases for the extreme values of *t*_*g*_. For all of the possible values of *t*_*g*_, and corresponding values of *R*_*0_e*_ and *p*, the reported case curves generated do not change much, for each of the countries, and lead to the same implication that total infection levels in the countries have grown to the point where new cases are declining due to insufficient numbers of susceptible people (for the current value of *R*_*0_e*_, i.e., under current disease control conditions).

The effective basic reproduction numbers, *R*_*0_e*_, in Table 1 range between 1.4 (in Bolivia) and 2.0 (in Madagascar). Estimates for the basic reproduction number, *R*_*0*_, the “natural” value in the absence of social distancing, for SAR-CoV-2 (the virus that causes COVID-19) in Wuhan at the outset of the global epidemic, range from 1.4 *[9]* up to 5.7 *[10]. R*_*0*_ might be expected to be higher in low-income countries due to factors such as dense living conditions, lack of access to clean water and sanitation facilities, and inability of most people to work from home. Thus, our findings suggest that, for the 10 LMICs studied, social distancing measures and practices, perhaps in combination with higher levels of partial immunity from prior exposure to coronaviruses *[11,12]*, likely reduced the effective basic reproduction number and slowed the spread of the disease – but with *R*_*0_e*_ above 1, they did not “crush the curve”.

Detection rates are estimated to be very low, ranging from 2.86% in Colombia to 0.05% in Malawi. These low detection rates explain how high levels of actual infections could be reached despite low numbers of reported cases, relative to total population, in all the countries studied. These low detection rates are not surprising. Serological testing results in Kenya, Pakistan and South Africa suggest that the number of people with coronavirus antibodies substantially exceeds the reported cases – by factors of up to 3,800 in Kenya (based on data in mid-May) *[13]*, of up to 540 in Pakistan (based on data from May to July) *[14]* and of up to 65 in South Africa (based on data from July to early August) *[15]*, which would correspond to detection rates of 0.03% for Kenya, 0.18% for Pakistan and 1.5% for South Africa. Furthermore, there is evidence that serological tests might underestimate effective immunity levels, because people may have partial immunity due to T-cell responses even if infection by SARS-CoV-2 did not produce antibodies or if those antibodies subsequently waned *[11,12]*.

In all these countries, the analysis indicates that significant percentages of their populations have been infected and have become immune – at least temporarily. The best estimates of total infection levels on 7 September derived from the fitted curves range from 45% in Bolivia to 79% in Madagascar *[16]*. Note that the infection levels required to curb the outbreak (the herd immunity threshold) normally quote the percentage of population infected at the peak of the curve, but significant numbers of people continue to be infected after this point, even as the numbers of new cases decline.

The infection fatality ratio (IFR), or the percentage of deaths from COVID-19 among those infected with the SARS-CoV-2 virus, can be estimated for countries with reliable estimates of deaths. For Bolivia, Colombia and South Africa *[17]*, the IFRs calculated from reported deaths divided by the total number of infections derived from our analysis, are 0.15%, 0.10% and 0.04%, respectively. In these three countries, estimates have been made of excess deaths due to natural causes, and, if all of these excess deaths are due to COVID-19, the IFRs for the three countries could be up to 0.57%, 0.13% and 0.11%, respectively *[18]*. All three countries are expected to have a lower overall IFR, compared to European countries, because their populations have a higher share of young people, who are significantly less likely to die from COVID-19 if they contract the virus. Differences in population age profiles explain the estimated IFR for Bolivia, but not for Colombia or South Africa. If reported infection fatality ratios by age, based on data from several HICs *[19]*, were valid for these countries, the expected overall IFRs for Bolivia, Colombia and South Africa would be 0.57%, 0.63% and 0.33%, respectively. Possible explanations for why mortality risk for COVID-19 might be lower in Colombia or South Africa, compared to the (mainly) European countries from which IFRs by age are derived, could include differences in immune-system response (already observed, for example, between men and women in some HICs), partial immunity to COVID-19 due to prior exposure to other coronaviruses, differences in lethality and prevalence of different virus strains, and different infection levels for different age groups.

To test the robustness of our approach, we applied the same methodology to fit curves to the first peaks in the comparison countries shown in Figure 3 (represented as red squares in this figure). Researchers at Imperial College London and others argued convincingly in June that European countries have not reached herd immunity *[20]*, and subsequent increases in cases have proven their point. Applying our model to fit curves just to the first peaks (shown with blue circles in Figure 3) *[21]*, and hence assuming that the peaks were due to herd immunity, we find that the best-fit curves match the observed data for the first peaks fairly well in all cases, but with values for the disease parameters that are implausible. For example, for France, the best-fitting curve yields *R*_*0_e*_ = 1.4, *t*_*g*_ = 3 days (the lowest permitted value) and *p* = 0.39%. The detection rate is well below the detection rates of between 7% and 18% suggested by serological studies in European countries *[22]*. New Zealand is well-known for “crushing” the curve – and the parameters associated with the best-fit “herd immunity curve” to its reported cases would be *R*_*0_e*_ = 1.9, *t*_*g*_ = 3 days and a highly improbable *p* = 0.03%. Thus, our approach leads to a conclusion for the comparison countries that the evolution of reported cases was <u>not</u> due to herd immunity (but instead must have been due to control measures). With this check, we increase our confidence in the hypothesis that the outbreaks in the 10 LMICs studied <u>are</u> declining due to herd immunity, which generates well-fitting curves with plausible parameters.

## Discussion

Prominent models of the epidemic from teams at Imperial College London (ICL) *[23]* and the University of Washington Institute for Health Metrics and Evaluation (IHME) *[24]* use SEIR simulations and determine key parameters – especially the effective reproduction number, which can vary over time – by fitting the models’ results for deaths to the reported numbers of deaths from COVID-19 and utilising age-specific IFRs from recent studies (in HICs). These models estimate that total infections to date for the 10 LMICs we studied are much greater than reported, but much smaller than our analysis suggests. For example, the models estimated total infection levels for South Africa of 7.7% (ICL) and 9.5% (IHME), and corresponding case detection rates of 13.9% and 11.2%, as of early September *[25]* – implying infection levels significantly below the levels of up to 40% suggested by serological study findings in Cape Town from July to early August *[15]*. It has previously been observed, by the IHME COVID-19 Model Comparison Team, that the predictive performance of seven COVID-19 models, including those of ICL and IHME, shows significantly higher errors for Sub-Saharan Africa, South Asia and Latin America and the Caribbean, compared to their performance for HICs *[26]*. Our research suggests why this might be the case. The ICL, IHME and other models are well-suited to HICs: reported deaths for such countries are likely to be reasonably close to actual deaths; the IFRs used in the models are based mainly on studies conducted in HICs; and it is clear from the evolution of reported cases that these countries have not reached herd immunity *[20]* and that their effective basic reproduction numbers have varied significantly over time as disease control measures have been introduced and adjusted (necessitating the additional granularity of SEIR modelling). However, for some LMICs: reported deaths from COVID-19 are likely to understate actual deaths by large factors *[18,26]*; age-specific IFRs might differ substantially from those in Europe and North America; and a simple model, using the approximation that effective basic reproduction numbers and detection rates remain constant over time, may be sufficient to describe the evolution of reported cases well (at least for the 10 countries we studied).

Systematic studies of representative population samples should be conducted in the 10 LMICs discussed here (and perhaps other countries as well) to determine the percentages of people who have been infected and are immune – and consequently test directly the primary conclusion that the overall infection levels are high and likely exceed effective herd immunity thresholds.

Even when susceptibility levels are sufficiently low that the virus can no longer spread exponentially, it will not be gone completely. Individuals may still contract the virus if they are not immune. Isolated communities, such as rural areas far from urban centres, may have much lower infection levels than the overall population, and may still experience localised outbreaks. New general outbreaks might happen as control measures are relaxed, and such outbreaks could be large for countries where *R*_*0_e*_ is currently close to 1 and/or for which cases have not mostly declined from the current peak – because relaxing disease control measures will increase *R*_*0_e*_ and could shift the effective herd immunity threshold to new values that are substantially greater than the current share of the population with immunity.

It is not yet known how long immunity from SARS-CoV-2 lasts. Even if a population is protected due to a high immunity level today, it is possible that this could be lost over time, which might lead to future outbreaks – the severity of which would depend on the share of people losing immunity, the amount of variation in timing of when people lose immunity, and whether susceptibility to reinfection is equal to the susceptibility to first infection. New strains of SARS-CoV-2 have emerged, and it is conceivable that future mutations could allow the virus to evade immune systems, and thus render previously immune populations susceptible again to the disease – but there is no evidence yet of any such immunity-evading strains.

## Supporting information

Supplementary materials including methods

## Data Availability

All data used is publicly available data (as published by WHO). Relevant sources are included in footnotes where applicable. All equations and methods used in this article are described in the supplementary material which we submit at the same time as a separate publication.

https://covid19.who.int/?gclid=Cj0KCQjwqrb7BRDlARIsACwGad70vhvBpGrU5OzlW0YlQIWvqdC5JdvmP6jSYm14cVkMyRsMnQvtILIaAhwcEALw_wcB

## Acknowledgements

We wish to thank Muhannad Alramlawi and Yohann Sequeira for help with parts of the modelling and research. We also acknowledge many useful discussions with Partners and consultants at Dalberg Advisors, in particular Edwin Macharia.

## Author contributions

A.S.L. conceived the first analysis of South Africa data, and J.P.C. conceived the idea to study a range of LMICs and comparator countries. A.S.L., C.J.A.N. and O.F. undertook the modelling. J.P.C. conducted the comparison of model parameters and assumptions with other research and with other epidemiological models. J.P.C. and C.J.A.N. prepared the manuscript.

## Data and materials availability

All data used in this study can be freely downloaded from the cited sources. The supplementary materials provide additional details on methods, including the equations used and the process for determining the parameters, and other notes and observations.

## Supplementary Materials

A separate document provides supplementary materials, including methods and other notes.

