## Supplementary materials including methods for "Evolution of COVID-19 cases in selected low- and middle-income countries: have they peaked due to high levels of infection and immunity?"

*Axel S. Lexmond[1]*, Carlijn J.A. Nouwen[2]*†, Othmane Fourtassi[2] and J. Paul Callan[2]*†*

*[1] Department of Engineering, University of Pretoria, South Africa*
*[2] Personal capacity*
** These authors contributed equally to this work.*

*25 September 2020; revised 1 October 2020 and 12 October 2020*

This paper contains the supplementary materials to the article "Evolution of COVID-19 cases in selected low- and middle-income countries: have they peak due to high levels of infection and immunity?" by the same authors. In that article, we examined the outbreak curves for 10 selected low- and middle-income countries (LMICs) and showed why the hypothesis that in these countries, COVID-19 cases have declined mainly through low susceptibility levels, is an option that warrants serious consideration. A simple model based on that hypothesis (a linearised version of Reed-Frost), with only a few constant parameters, fits the observed case data remarkably well, and yields parameter values that are reasonable.

In this paper, we describe the method we have used to determine the best-fitting curve for each of these countries. We use the case of South Africa to provide the full analytical detail. Subsequently, we explain how we apply this method to the other 9 countries.

### 1. Model and equations used to describe the outbreak to date

As we want to test the hypothesis whether the outbreak curve in these countries can be described a peak driven by low susceptibility, we use a set of equations that define a 'natural' exponential outbreak. For this, we use a homogeneous population model which describes a basic exponential growth outbreak model with constant parameters. In this model, we assume we can describe the outbreak in cycles. The number of people that is infected in cycle i+1 is given by

eq. 1) $n_{i+1} = n_i R_{0\_e} \left(1 - \frac{n_{immune}}{n_{tot}}\right)$

Where

- $n_i$ is the number of people that have been infected in infection cycle i
- $R_{0\_e}$ is the reproduction number of the virus, taking into account any measures and behaviour change but not taking into account any immunity. We call this the 'effective *basic* reproduction number' to distinguish it from the effective reproduction number $R_e$[1]
- $n_{tot}$ is the total population
- $n_{immune}$ is the average number of immune people during an infection cycle (see section 8 for averaging method)
- and $R_{0\_e} \left(1 - \frac{n_{immune}}{n_{tot}}\right)$= the effective reproduction number, $R_e$. This is the number of people any one infected person at a particular moment will infect on average, taking into account both behaviour and the build-up of immunity in the population.

Assuming everyone who has been infected becomes immune, the number of immune people at the beginning of the cycle from i to i+1 is given by: $n_{immune}(i) = \sum_{j=0}^{i} n_j$.

The number of people reported over the course of infection cycle i to be newly diagnosed with COVID-19 is given by:

eq. 2) $x_i=pn_i$ where p is the detection rate

The infection curve started when the first infected person entered South Africa ($n_0$=1). To express the number of cases as a function of time (t, in days) rather than as a function of the number of infection cycles, we use equation 3:

[1] Three distinct reproduction numbers characterise the disease in different situations. First, the basic reproduction number, $R_0$, is the average number of new infections caused by each current infected individual, that would happen in the absence of any disease control measures by governments or individuals, and before there is widespread immunity. Even this number is not a universal characteristic, because it can vary by location, even in the absence of control measures, due to differences in typical numbers of social contacts in different places. Second, we define the effective basic reproduction number, $R_{0\_e}$, as the adjusted basic reproduction number after control measures are taken, but without significant levels of immunity. This number can, therefore, change from time to time, as governments change policies and people change their practices. Third, the effective reproduction number, $R_e$, is the actual average number of new infections caused by each current infected individual, which decreases as the number of people with immunity increases (and also depends on disease control policies and practices).

eq 3) $t=t_g*i-\Delta t$ where t = time in days since first reported case

The model has 5 parameters:

- $t_g$ generation time – which is the cycle time with which we run the model[2]
- $R_{0\_e}$ effective basic reproduction number of SARS-CoV-2
- $n_{tot}$ total population
- $\Delta t$ number of days that the SARS-CoV-2 has been spreading undetected
- p detection rate

The model has one dependant (output) variable

- $x_i$ The number of reported people with COVID-19

Of the 5 independent variables, 1 variable is constant ($\Delta t$) and 2 other variables are most likely constant as well (particularly for the timeline we are considering): the generation time and the total population. That gives 2 independent variables that could change over time: the effective basic reproduction number of the virus ($R_{0\_e}$) and the detection rate (p).

**Our model is a linearised version of the classic Reed-Frost model, a simple deterministic model for simulating the spread of a disease outbreak.** The formula for the Reed-Frost model is:

eq. 4) $n_{i+1} = s_i(1-(1-q)^{n_i})$

where:

- $n_i$ is the number of people that have been infected in infection cycle i
- $s_i$ is the number of susceptible people after infection cycle i (which equals the total population minus the sum of all those infected up to and including that cycle)
- q is the probability that, within one time period, an infected person will come into contact with any other person in the population and will transmit the infection to that other person.

In this model, the effective basic reproduction number, the average number of people to whom the disease is transmitted by one infected person, in one infection cycle, when the entire population is susceptible, is simply:

eq. 5) $R_{0\_e} = n_{tot}\, q$

where $n_{tot}$ is the total population.

The duration of one infection cycle in this model is the generation time ($t_g$), which is the average time from infection of one person and when that person infects other people.

---

[2] Understood as follows: if person Y infects multiple people, this is unlikely to happen all at the exact same time. Hence, one determines the time at which Y got infected to the moment Y infects the other person for each of the people Y infects. Averaging this number across all the people Y infects, gives the equivalent of the infection cycle for Y. To get to this number for *the virus* ($t_g$ as used here), this 'individual' infection cycle is averaged across all people who infected someone. At population level, $t_g$ is likely to *decrease* as the degree to which symptomatic patients self-isolate increases, and likely to *increase* as the proportion of asymptomatic cases increases.

The linear approximation of the Reed-Frost equation (4) is:

$$n_{i+1} \approx s_i \left(1 - (1 - n_i\, q + \cdots)\right) = s_i\, n_i\, q$$

which, substituting for $R_{0\_e}$ using eq. (5) and for $s_i = n_{tot} - n_{immune}$, yields:

$$n_{i+1} \approx n_i \frac{R_{0\_e}}{n_{tot}} \left(n_{tot} - n_{immune}\right)$$

This is the same as eq. (1) above.

Our model can also be described as a compartmentalised SIR model with constant parameters. In SIR models, people are either susceptible, infected or have recovered. We model the “exposed” (infected-but-not-yet-infectious) section of the population by running the outbreak through “infection cycles”. The duration of an infection cycle, the generation time, is the average time of the “exposed” stage in more complex SEIR models. The equations used describe the process in the same way as the differential equations used in SEIR models yet we use discrete time periods (infection cycles) rather than dt.

### 2. Selection of a country to illustrate fitting the parameters with

We identified 10 low- and middle-income countries (LMICs) for which the observed pattern in the actual COVID-19 case numbers might be explained assuming cases peaked and declined as a result of low susceptibility. Of these 10 countries, on first visual inspection, South Africa seems to have the best fit. Figure 1 shows the observed data (the circles) and the best fitting curve (result of the process as described in this article) using a standard exponential outbreak curve (the red line), visually showing a good fit. South Africa also has a relatively large number of datapoints after the peak which makes the fitting procedure more accurate and easier to illustrate than when applied to countries which hit their peak more recently. In the remainder of this document, we will *illustrate* the application of the approach using South African data and references. In section 5, we will discuss the process of applying this approach to other countries – as well as the implications of poor data quality and the associated fitting approach in that case.

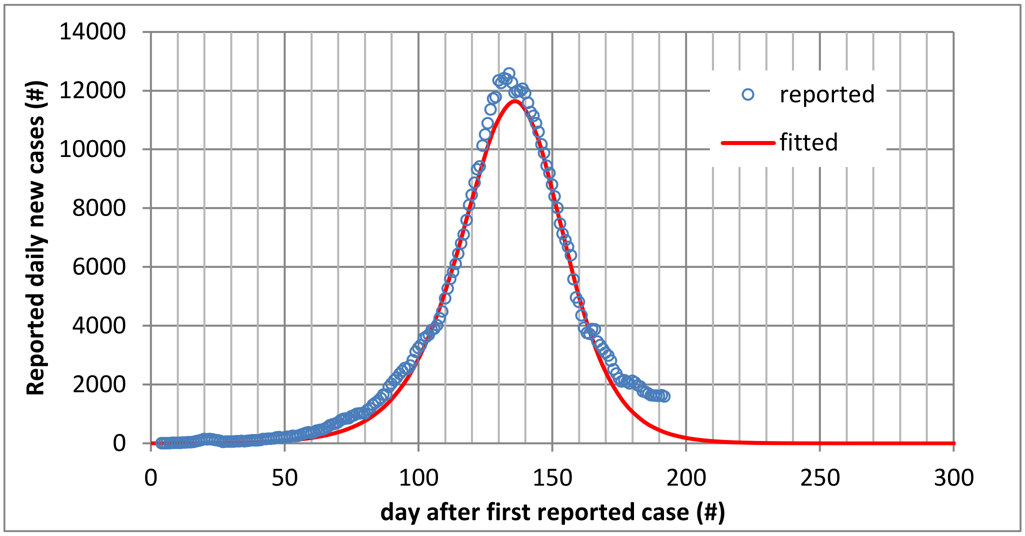

*Figure 1: Daily cases of COVID-19 in South Africa over time*

### 3. The method used to derive the parameters

#### *Each parameter impacts the shape of the outbreak curve in a different way*

As outlined above, three of the five variables can be assumed to be constant over the period we are considering (and one of those, the total population of a country, is a known entity). Interestingly, each of the variables have a different effect on the shape of the outbreak curve:

- Three of the four variables have a linear transformation effect on the curve (meaning they do not fundamentally alter the *shape* of the curve)
    - Δt shifts the entire curve horizontally (to 'later' or 'earlier') without altering the shape of the curve
    - p stretches the curve vertically (basically changing the scale on the vertical axis) without fundamentally altering the shape of the curve
    - $t_g$ stretches the curve horizontally (changing the scale on the horizontal axis). Whilst this is a linear transformation of the curve and does not fundamentally alter the shape of the curve the way $R_{0\_e}$ does, $t_g$'s effect is the hardest to separate out from $R_{0\_e}$
- Only $R_{0\_e}$ fundamentally alters the shape of the curve: $R_{0\_e}$ defines the ratio between the growth rate during the increase of the daily caseload and the negative growth rate during the decrease of the daily caseload. This effect is illustrated in Figure 2 below. For a fixed doubling time (which, as discussed later, is a function of both $R_{0\_e}$ and $t_g$) assumed to be 10 days in this figure, this graph provides curves for daily caseload numbers (normalised to a scale from 0 to 1 on log-lin scale) for different levels of $R_{0\_e}$. The higher $R_{0\_e}$ is, the greater the ratio between the pace of decline and the pace of growth – and the more 'skewed' the curves become.

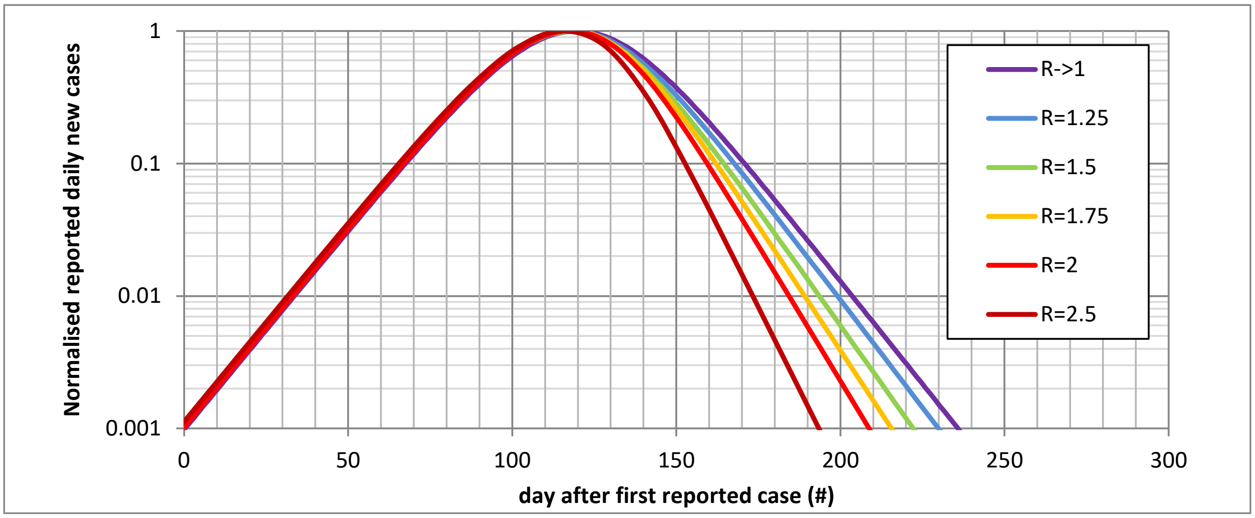

*Figure 2: Normalised outbreak curves (log-lin scale) for a doubling time of 10 days and various values of $R_{0\_e}$ showing how $R_{0\_e}$ changes the 'shape' of the outbreak curve*

#### *The benefits of an analytical approach: covariance between $R_{0\_e}$ and $t_g$ limits value of multi-parameter regression*

$R_{0\_e}$ and $t_g$ are inherently linked. From the exponential growth rate in the early part of an outbreak, it follows[3] that the doubling time equals $\frac{\ln 2}{\ln R_{0\_e}} t_g$

This link introduces the complexity that it is hard to determine the exact unique combination of $R_{0\_e}$ and $t_g$ that leads to the observed curve; there often are a range of combinations of $R_{0\_e}$ and $t_g$ which can produce very similar curves and thus have a very similar goodness-of-fit.

We illustrate this in the figures below for South Africa. Figure 3 provides the link between $R_{0\_e}$ and $t_g$ – for a broad range of of $t_g$ – from 3 to 11 days – in increments of 1 day, we identified the value for $R_{0\_e}$ that yields the best possible fit (using least squares method).

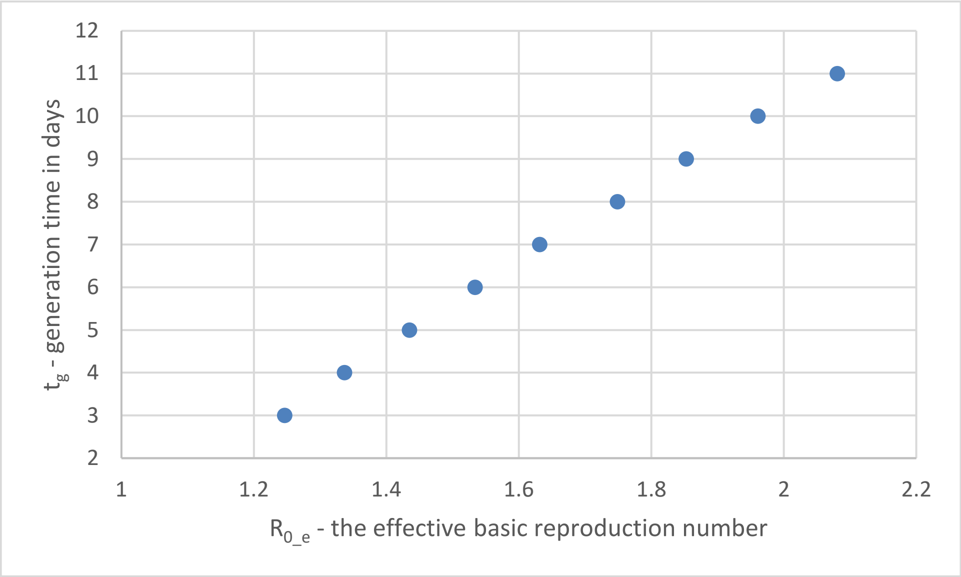

*Figure 3: Relationship between $R_{0\_e}$ and $t_g$ (combinations yielding the curve with the best fit) for South Africa*

Figure 4 provides the associated value of $R^2$ – the goodness-of-fit measure using the least squares method for each of these datapoints. $R^2$ is very high for all, indicating that the curve generated with these parameters provides a very good fit for the observed data. Importantly, though, $R^2$ hardly varies over a wide range of parameters: over the full range of $t_g$ from 3 to 11 days and the associated range of $R_{0\_e}$ from 1.2 to 2.1, the $R^2$ is 0.99. What this shows, is that because of the strong covariance between $t_g$ and $R_{0\_e}$ it is impossible to determine the exact unique combination that leads to the observed curve. The wide range of underlying values introduces an uncertainty that is not particularly helpful – which is why we sought an analytical approach to deriving the parameters from the observed caseload data. Once one of them is determined, the relationship between them makes it possible to determine the other one. We then go from a fitting exercise with 3 variables to one with only 1 variable left: the

[3] See section 9 for full derivation of the equation

detection rate (p), which – as we shall see – can also be derived analytically provided that $R_{0\_e}$ has been defined analytically and the outbreak has 'passed the peak'.

Note that this very issue of strong covariance is why it is very hard to determine the basic reproduction number, $R_0$, for any new disease, even when the case doubling time (as determined by $\frac{\ln 2}{\ln R_{0\_e}} t_g$ ) is well-known. The equations we use, assume $R_{0\_e}$, p and $t_g$ are constant during the entire outbreak. Once we have determined the parameters, we will test this assumption by considering whether the resulting fitted curve fits the observed caseload well with reasonable assumptions for the parameters. In section 6, we will discuss these assumptions in more detail.

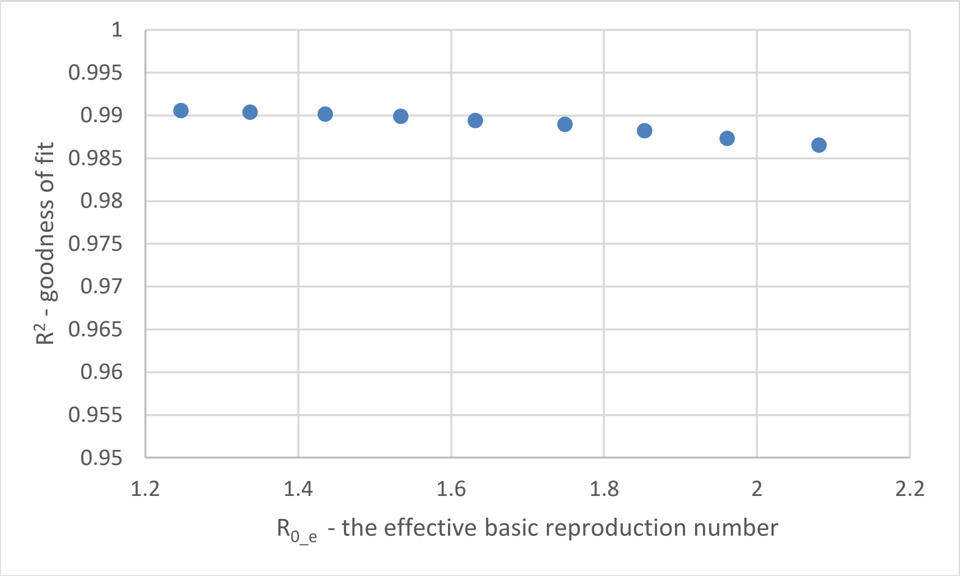

*Figure 4: Value of $R^2$ (goodness of fit) for full range of combinations of $R_{0\_e}$ and $t_g$ for South Africa*

### *A simple analytical method to derive $R_{0\_e}$ and $t_g$*

#### Derivation of $R_{0\_e}$

The number of daily new cases grows exponentially well before the peak, and decreases exponentially well after the peak. The growth and decline are functions of both $R_{0\_e}$ and $t_g$, but the *ratio* of growth and decline is *only* determined by $R_{0\_e}$.

Exponential growth and decline: $n = C * 10^{(t-t_0)/\tau}$

where C is a constant.

$$\log_{10}\left(\frac{n_2}{n_1}\right) = \frac{t_2 - t_1}{\tau}$$

$$\tau = \frac{t_2 - t_1}{\log_{10}\left(\frac{n_2}{n_1}\right)}$$

Where $t_1$ and $t_2$ are 2 moments during incline (for $\tau\uparrow$) or decline (for $\tau\downarrow$)

$\tau\uparrow$= time for cases to incease ten-fold (well before peak of the outbreak)

$\tau\downarrow$= time for cases to decease ten-fold (well after peak of the outbreak)

Figure 5 below plots the daily caseload on a log-lin scale (for just a part of the outbreak, in order to better see the relevant section). In this figure, the red lines show the inclines of both the growth and the decline in the caseload – each of which are exponential processes. As articulated above in the equations, the *ratio* between the growth and decline exponents is *solely* a function of $R_{0\_e}$. These red lines are chosen as follows:

- Firstly, we want to determine the incline at portions of the curve that are both straightest on log scale (indicating that both p and $R_{0\_e}$ can be assumed constant) and within the lockdown period (to have relatively consistent circumstances).
- For the decline period, we do not have a long set of data to choose from – so we took the straightest final part of the curve (the further away from the peak, the straighter the line is on log-lin scale).
- We are interested in the ratio; and the shape of the curve to either side of the peak is very comparable (although the inclines will be different, which is linked to $R_{0\_e}$, the *shape* to either side of the peak is very similar which is a mathematical result).
- Therefore, to get the most accurate estimate of the ratio, we should take the incline pre-peak that covers the same number of daily cases as the period chosen to the right of the peak does (in this case ranging from 3,500 to 8,500 cases per day). We could also have taken the much longer stretch to the left of the peak; that would create greater accuracy for the incline but reduce comparability with the decline portion.
- This logic makes us determine incline and decline over the red periods in Figure 5 below.

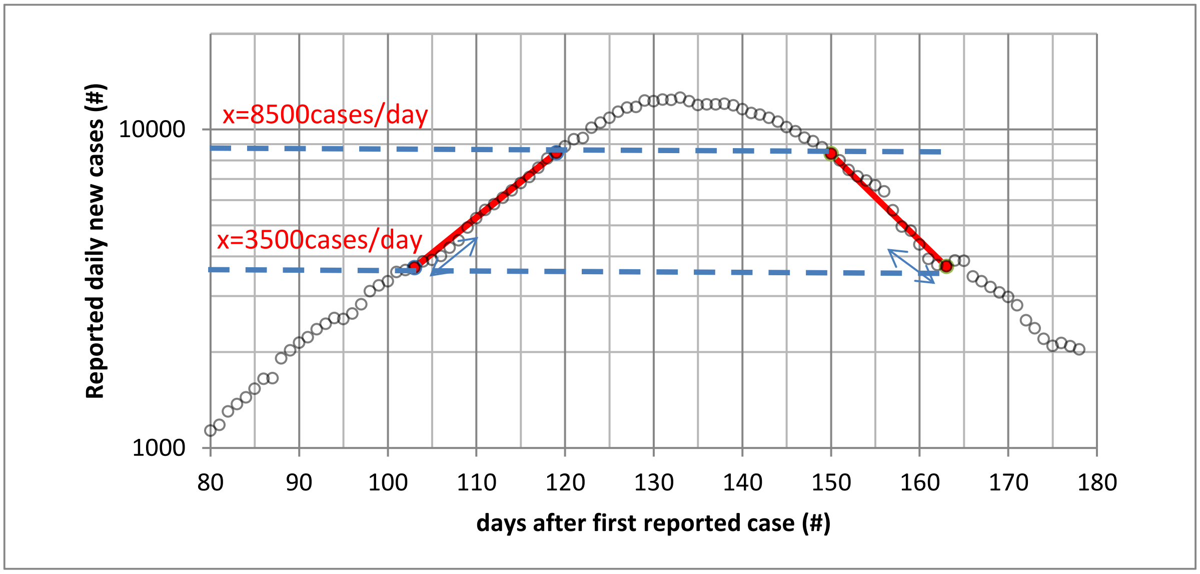

*Figure 5: Selected interval range for determination of ratio of incline and decline - to analytically determine $R_{0\_e}$*

In order to increase accuracy, we calculated the inclince and decline and their ratio a number of times (indicated by the blue double-headed arrows in the figure above). In all cases, we kept the highest point fixed (as above that, very quickly the curve deviates from being a straight line). We then varied the lowest point to get various intervals and applied a regression analysis on the actual measured caseload data to determine the value of τ↑, and τ↓. As the curve is less steep before the peak, the same stretch on the vertical axis incorporates more datapoints than to the right of the peak.

- Thus, to determine the value of τ↑,we were able to use 10 intervals for which a reasonably ‘matching’ pair could be found in the decline[4]: each ending at day 119 and start date ranging from day 101 up and until day 109.
- To determine the value τ↓, we could use 5 intervals: each starting at day 150 and end date ranging from day 159 to day 163

The regression outcomes for each of these intervals are provided in Figure 6.

[4] At first, we used 15 intervals during the period of incline but for 5 of them, no reasonably matching pair could be made with a decline interval – using these would have introduced too much discretion and potentially inaccuracy

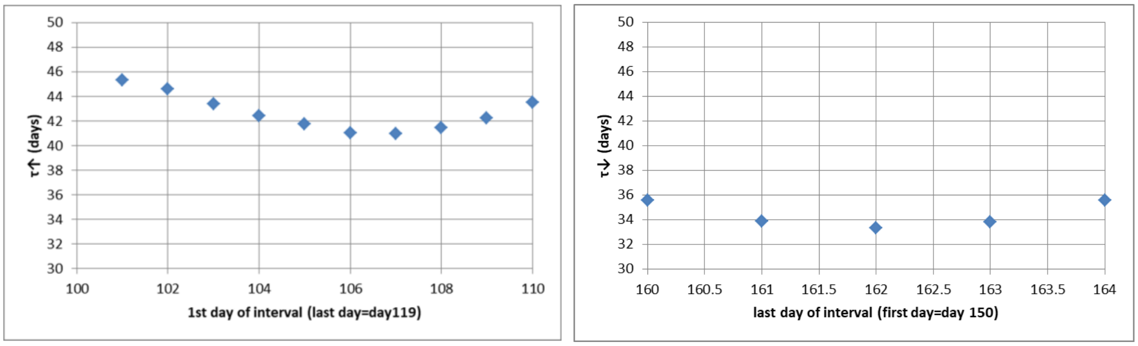

*Figure 6: Regression outcomes for τ↑ for each of the 10 intervals used (left) and for τ↓ for each of the 5 intervals used (right)*

We then combine these datapoints to find 'matching' pairs – see Figure 7. This requires some judgment as we have only 5 values for τ↓ and 15 for τ↑. We matched the datapoints (with weighted averaging and intrapolation) to get 10 pairs and thus 10 ratios.

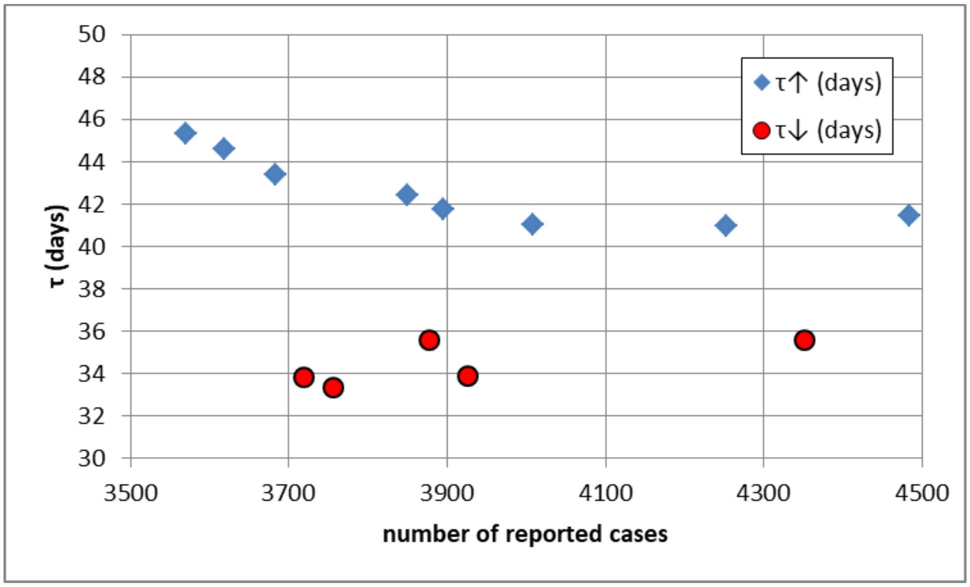

*Figure 7: Pairs for estimates of incline ratio - x axis is number of reported cases at lower end of interval*

This provided a range for the τ↓/ τ↑ ratio for each of the 10 pairs. Figure 8 shows the relationship between this ratio and the $R_{0\_e}$ using the equations in the model (the blue line). From this, we can derive the range for $R_{0\_e}$ for the range for the τ↓/ τ↑ratio which was derived from the reported case data (1.15 – 1.31).[5] This gives a range for $R_{0\_e}$ of 1.45 – 1.90. Averaging across the 10 resulting values for $R_{0\_e}$ (associated with the 10 ratios - thus not simply taking the average of the range) gives the **best-estimate $R_{0\_e}$ of 1.74**.

[5] Note that this method of using multiple intervals, was applied in order to deal with any errors in underlying data. Therefore, we do not provide a wider confidence interval on top of this range.

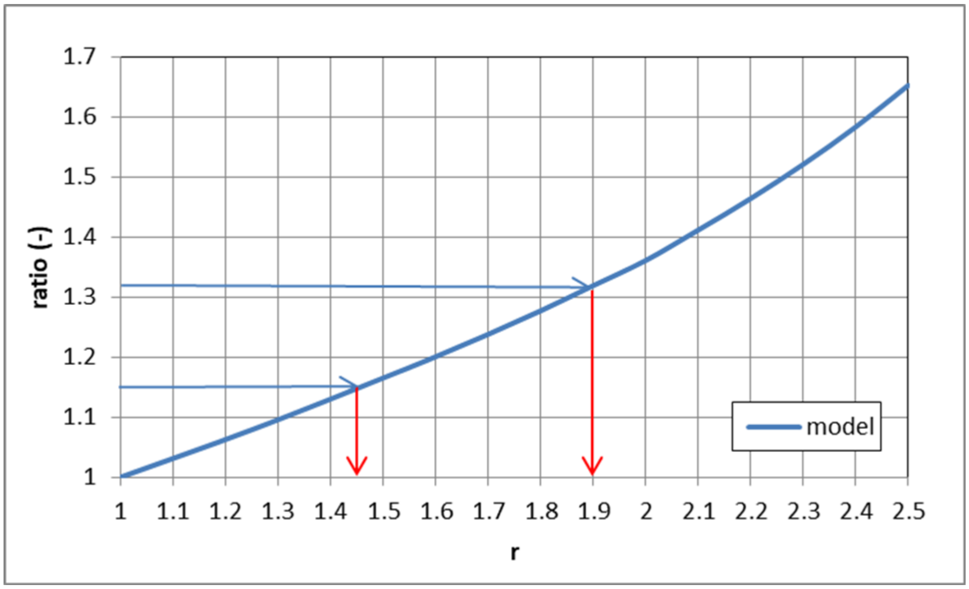

| range | τ↑ (days) | τ↓ (days) | τ↑ /τ↓ | R |
|---|---|---|---|---|
| highest ratio in range | 44.4 | 33.8 | 1.31 | 1.90 |
| lowest ratio in range | 41.0 | 35.6 | 1.15 | 1.45 |
| best estimate | | | | 1.74 |

*Figure 8: Range for incline ratios and $R_{0\_e}$*

#### Derivation of $t_g$

The width of a peak (W), defined as the time between the days where the daily number of infected persons is exactly half the maximum of the daily number of infected persons (so half the peak), depends on $R_{0\_e}$ and $t_g$ only. Since $R_{0\_e}$ is known and the width is a linear function of $t_g$, this allows one to determine $t_g$. This method is visualised in Figure 9 and captured in the equation below.

$$W = A\,(R_{0\_e}) \cdot t_g$$

where W = width of the peak and A is a constant. A is a function of $R_{0\_e}$ - and as $R_{0\_e}$ is constant, it follows that A is constant.

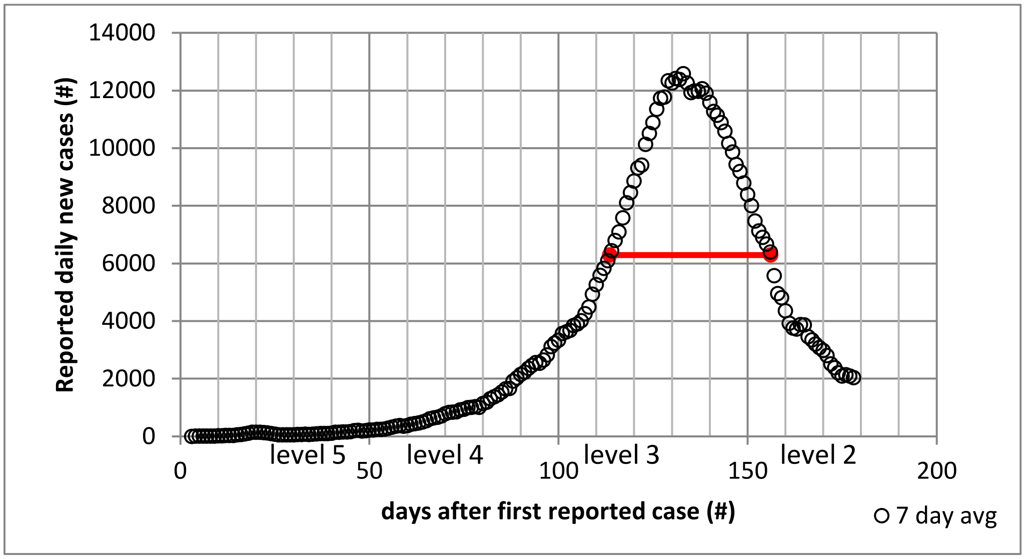

*Figure 9: Visualisation of the method to determine $t_g$*

First, we determine the width from the observed data. The maximum number of cases (7 day average) in the observed data is 12584 cases/day. Half this amount of cases is found by interpolation at 113.56 (left side of curve) and 156.13 (right side of the curve) days since the first reported case, so the width of the curve, defined in this way, is 42.6 days.

Next, we determine the value for $t_g$ as a parameter in the equation above such that we get the same width. A($R_{0\_e}$=1.74) can be determined from the simulated outbreak results, using the governing equations 1-3. For $t_g$ =10 and $R_{0\_e}$=1.74, W=54.5. From this it follows that A($R_{0\_e}$)=5.45. Filling in W=42.6 days and A=5.45 gives the generation time ($t_g$): $t_g$ **=42.6/5.45=7.8 days.**

For $R_{0\_e}$= 1.45, the associated $t_g$= 4.8 days and for $R_{0\_e}$= 1.90, the associated $t_g$= 9.1 days.

#### Derivation of p (the detection rate)

At the top of the outbreak, when the infection level is so high that the number of cases peaks and starts to decline, the number of infected people is given by $N=N_{tot}\cdot(1-1/R)$. The total amount of reported positive cases can also be determined from the outbreak data: X=Σx where X is the number of reported cases before the peak and x is the daily number of reported cases. The detection ratio is given by: p=X/N.

R ranges from 1.45 to 1.90, with a best estimate of 1.74. As a result, the estimated total number of infected people at the peak of the outbreak (day 137, N in the equation) ranges from 18.41 million (for R=1.45) to 28.09 Million for R=1.9, with a best estimate of 25.22 Million for R=1.74.

Best estimate for p (for R=1.74) p = 355,633 / 25,223,236 =1.41%.

High estimate for p: p=X/N=355,633 / 18,406,145 =1.93% (R=1.45).

Low estimate for p: p=X/N=355,633 / 28,093,590 =1.27% (R=1.9)

The resulting best-fitting curve has an associated $R^2$ of 0.984.[6] Visually, one can understand the 'fit' by seeing how well the model (the solid red line in Figure 10) overlays with the observed caseload (the circles).

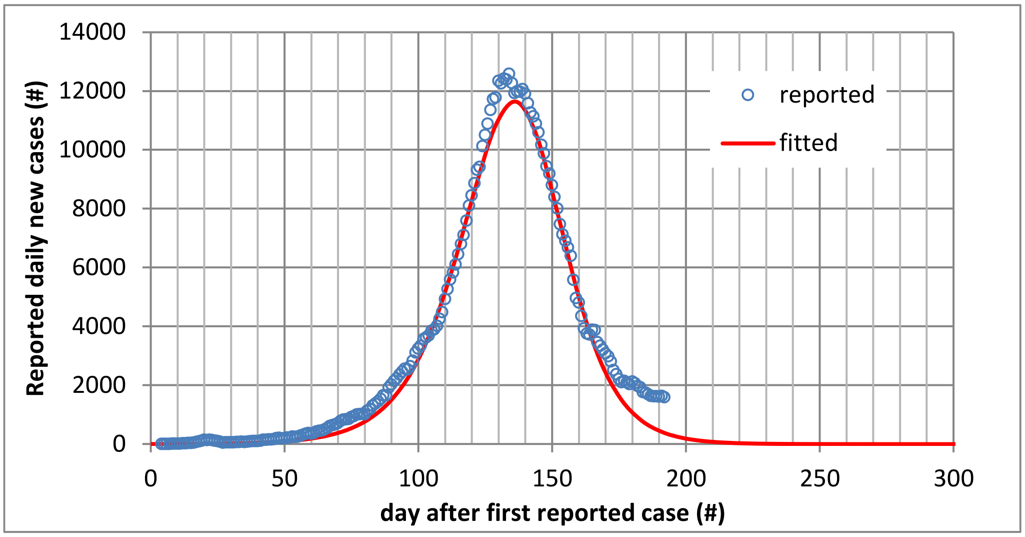

*Figure 10: Daily cases of Covid 19 is SA vs time. Model: $R_{0\_e}$=1.74, $t_g$ =7.8 days, p=1.41%; $R^2$ = 0.98.*

The model with constant parameters fits the data well – in other words: the entire pattern of observed daily cases can be explained with this particular set of parameters[7] that belong to a set of equations that describe an outbreak pattern that reaches a single peak driven by low susceptibility due to high levels of infection and immunity.

[6] This $R^2$ is marginally lower than the R2 for fits using the optimising algorithm. The analytical method focuses on data related to the core of the outbreak, using half-way width and incline/ decline ratios. The $R^2$ uses all data – so the values the optimising algorithm finds, will fit the entire curve, including the tail, a bit better than the associated $R^2$ (across the entire curve) for the analytically derived values.

[7] More complex modelling should be used if p and $R_{0\_e}$ cannot be assumed constant for a sufficiently long portion of the outbreak. However, for South Africa, the high $R^2$ now confirms that this assumption of constant p and $R_{0\_e}$ was a decent assumption.

### 4. What else could it be, if not low susceptibility due to high levels of infection and immunity?

We have posed the hypothesis that the South African case pattern can be explained by high levels of infection and immunity – and have shown that the best-fitting curve using constant values for p, $R_{0\_e}$ and $t_g$, has a strong fit with an $R^2$ of 0.99. But is it possible that the caseload goes down as observed in the *absence* of such high levels of infection and immunity? That is possible, if we do not assume $R_{0\_e}$, p and $t_g$ to be constant. Note that the pattern of observed number of cases (the circles in Figure 1) can be reproduced by solving eq. 1-3 while varying $R_{0\_e}$ as a function of time (assuming a constant p) OR by varying p at constant $R_{0\_e}$. We have already seen that the *shape* of this type of a curve is uniquely determined by $R_{0\_e}$. Yet, we did assume that both p and $R_{0\_e}$ were constant over a sufficiently long, relevant part of the outbreak that we use to determine the parameters. We will now show what alternative explanations would look like focusing on stress testing that particular assumption. We will show why alternative explanations are much less likely than accepting the hypothesis of low susceptibility due to high levels of infection and immunity.

Firstly, the reported number of cases can go down in the absence of sufficiently low susceptibility[8] because the **detection rate (p)** has gone down: i.e., the testing strategy has changed and as a result, a greater proportion of cases go undetected. That would lead to a lower reported number of cases whilst the real number of cases continues to increase (because we have not reached a sufficiently high level of infection and immunity). *However,* a once-off drop in detection rate in the absence of sufficiently low susceptibility would result in a once-off drop in the number of cases – after which exponential growth continues. Figure 11 shows what this would look like: the red and blue lines show this scenario with red representing the caseload observed before the drop in detection rate and blue after the once-off drop. These curves combined look nothing like the actual observed case numbers (the circles). So a once-off reduction in detection rate cannot have driven the reduction in observed cases.[9]

[8] By 'sufficiently low' susceptibility – or conversely 'sufficiently high levels of infection and immunity' we mean levels level at which the remaining susceptible population is not large enough to sustain further growth of the epidemic, thus causing a peak and subsequent decline in cases.

[9] Interestingly, the number of confirmed cases DID show exactly this type of discontinuity *on the exact day the lockdown started*. As any change in $R_{0\_e}$ due to lockdown would not show instantaneously (but at least have a ~7 day timelag), this points towards the fact that testing approach changed as lockdown started – and became overall less effective. There is at least anecdotal evidence for this statement: early on during the lockdown, entire (rural) communities were tested even if no cases had been present in them to understand spread to date. Conversely, before the lockdown, focus was more on track-and-trace of individual cases.

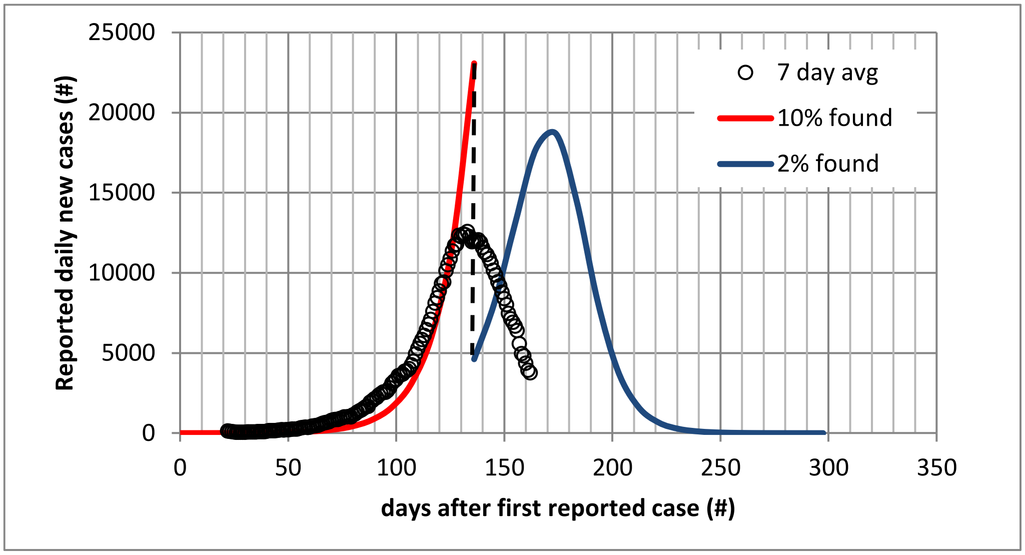

*Figure 11: Scenario with a sudden change in testing approach and thus detection rate*

A more *continuous* drop in detection rate, and not a change in $R_{0\_e}$ or peak due to low susceptibility, could also result in a reduction in number of observed cases. For that to result in the reduction as observed, the detection rate mid-September should be over a factor 1000 smaller than the detection rate at the start of level 5 lockdown. For this to materialise, the testing approach needs to change very dramatically (to result in a factor of 1000 poorer detection result) yet gradually and continuously (to result in a smooth curve rather than discontinuity shown above). As the cases began to drop, the South African government examined the testing approach – they checked, for example, whether there was a shortage in reagents that could cause a drop in testing and thus a drop in the reported number of cases even if the real number of new cases was increasing or stable. They explicitly communicated this was not the case – making a deterioration of a factor of 1000 an extremely unlikely alternative explanation for the drop in number of cases.

Another potential explanation for a drop in case numbers, is a **reduction in the effective basic reproduction number (**$R_{0\_e}$**).** In Figure 12 below  the straight line on a log-lin scale shows that the exponential growth rate was largely constant for a long period of time (between day 30 and 120 since first confirmed case).

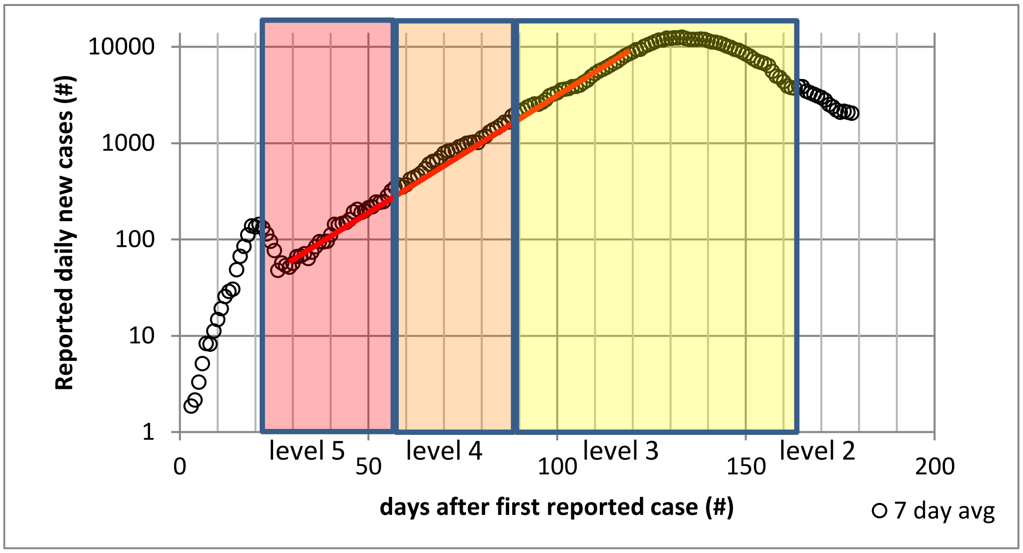

*Figure 12: Daily cases in South Africa on a log-lin scale*

When we zoom in, in Figure 13, we show that this is not *exactly* right: the curve slopes slightly downward. If we attribute this slight deviation from the straight line from Figure 12 fully to a change in $R_{0\_e}$ (and thus assume p constant), $R_{0\_e}$ would have had to decrease by 22% over a period of 90 days to drive the slight downward deviation from the straight line.[10]

[10] This figure uses $t_g$ =8 to determine the value of $R_{0\_e}$ – yet the decline in $R_{0\_e}$ (thus the 22% and 75%) is independent of the exact value of $R_{0\_e}$ (thus, the values on the vertical axis are irrelevant to the analysis here). At a different value for $t_g$, $R_{0\_e}$ would be different but the necessary change observed in $R_{0\_e}$ in order to fit the full curve (assuming p constant) would not shift.

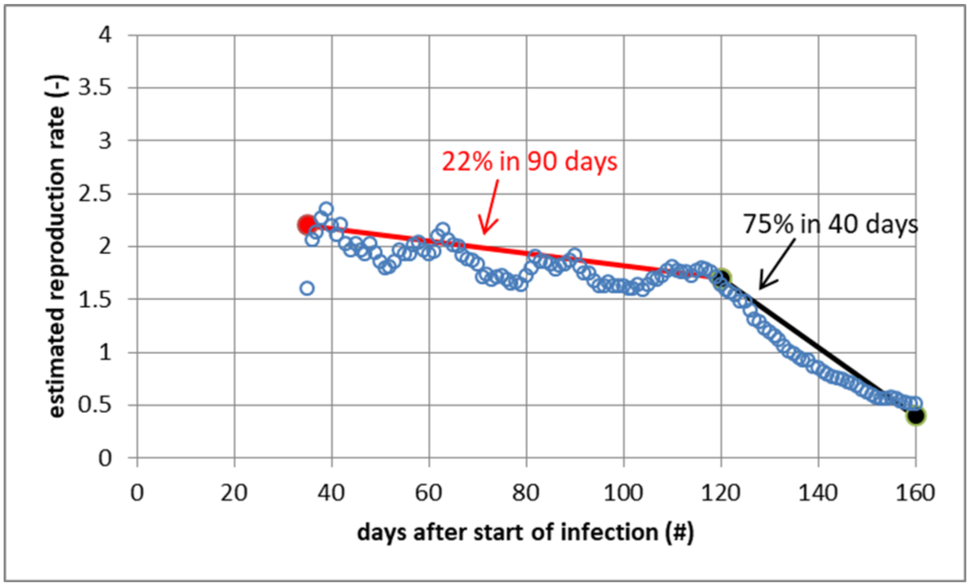

*Figure 13: Predicted $R_{0\_e}$ vs time (for constant p and $t_{g\ =\ 9.5\ days}$) What $R_{0\_e}$ should be over time, assuming a constant % of cases get detected, to get the evolution of the caseload as actually observed. Note that case numbers are a moving 14-day average to remove some of the empirical noise and note that the % is independent of $t_g$.*

The colour coding in Figure 12 shows the various levels of 'lockdown' that South Africa implemented: the darkest shade pink is the most stringent period, level 5, followed by level 4 and level 3. The exponential growth rate is a function of the virus reproductive process and of the behaviour of people: the more social distancing and contact restrictions, the lower the exponential growth rate is expected to be. Between days 30 and 120, South Africa saw a pretty substantial shift in its containment measures, going from strict lockdown level 5 to more lenient levels 4 and 3. One would expect the growth rate to increase (i.e., the growth to accelerate) as restrictions eased. Yet over that entire period, the growth was surprisingly constant and, if anything, decreased somewhat. However, in the middle of level 3 lockdown, the curve curbed strongly, suddenly and consistently. Figure 13 shows that if we tried to explain this entire shift through a shift in $R_{0\_e}$ (keeping p constant), $R_{0\_e}$ would need to start to decrease steadily – not once-off but continuously and in a very particular and rapid way – decreasing by 75% in ~40 days. That is not very realistic given how consistent the exponential growth rate (and, if we assume p constant, therefore $R_{0\_e}$) has been across various degrees of lockdown.

The most likely explanation for this pattern is thus not that the underlying $R_{0\_e}$ suddenly changed (and continued to do so in a specific pattern) when it had not over such a long period of time and range of circumstances. The most likely explanation is that infection levels has become so high that the remaining susceptible population is not large enough to sustain enable further growth.

### 5. Application of the methodology to other countries

#### Fitting of parameters for the 9 LMICs other than South Africa

In the absence of exact empirical determination of any of the parameters, it is impossible to determine the exact right value within this set. As we explain in our main paper, we assume the analytically derived value for $t_g$ of 7.8 days to be valid in other countries as well – yet we also present a credible range.

For the range, we varied $t_g$ in steps of 1 day each across a realistic spectrum from 3 to 11 days as explained in the main paper. We estimate $R_{0\_e}$, p and the horizontal shift delta t by fitting the curve optimally to the observed caseload data. This is done by minimising the sum of the squared residuals (and thus maximising the $R^2$), using a non-linear Generalised Reduced Gradient algorithm, iterating until the relative change in sum of the squared residuals is less than 10 ^ - 4 in each of the last five iterations. Subsequently, we run the optimisation again, starting with the resulting values of the previous run, until the relative change in the sum of squared residuals does not exceed 10^-4 for all of the runs done in an iteration of the algorithm. This process gives decent results yet is not perfect. It does not run a (near) infinite set of combinations with an unlimited number of decimals. Therefore, the parameter values resulting from this process still have an error margin. However, given the iterative process and the observation of really constant outcomes, this error margin is expected to be relatively low.

The resulting parameters, for $t_g$ of 7.8 days as a point estimate and a range from 3 to 11 days, are in *Table 1* below. Note that the values for South Africa are the analytically derived values (which associate with a range for $t_g$ of 4.8 to 9.1)

*Table 1: Key parameters for 10 selected LMICs for which we have shown that a decline in cases due to low susceptibility, is a very plausible hypothesis for their outbreak curve profile*

| **Country** | **Effective basic reproduction number ($R_{0\_e}$)** [see text] | **Detection rate (p)** | **% of population infected up to 7 Sept** | **R-squared for fitted curve** [perfect fit = 1] |
|---|---|---|---|---|
| Afghanistan | 1.7 (1.2 – 2.1) | 0.13% (0.26% - 0.12%) | 71% (37% - 82%) | 0.97 |
| Bolivia | 1.4 (1.1 - 1.6) | 2.35% (5.20% - 1.90%) | 45% (20% - 57%) | 0.99 |
| Central African Republic | 1.8 (1.3 - 2.2) | 0.13% (0.25% - 0.12%) | 74% (39% - 86%) | 0.94 |
| Colombia | 1.5 (1.2 – 1.7) | 2.86% (6.24% - 2.35%) | 46% (21% - 58%) | 1.00 |
| Egypt | 1.6 (1.2 – 1.9) | 0.15% (0.30% - 0.13%) | 64% (32% - 77%) | 0.98 |
| Kenya | 1.7 (1.2 – 2.0) | 0.09% (0.19% - 0.08%) | 66% (33% - 77%) | 0.98 |
| Madagascar | 2.0 (1.3 – 2.4) | 0.07% (0.12% - 0.06%) | 79% (44% - 89%) | 0.96 |
| Malawi | 1.6 (1.2 – 1.9) | 0.05% (0.09% - 0.04%) | 64% (32% - 77%) | 0.94 |
| Pakistan | 1.6 (1.2 – 2.0) | 0.20% (0.38% - 0.16%) | 66% (34% - 80%) | 0.95 |
| South Africa | 1.7 (1.5 – 1.9) | 1.41% (1.93% - 1.27%) | 71% (55% - 77%) | 0.98 |

### Fitting of parameters for comparison countries

In order to test whether our approach yields viable results across *too* broad a range of countries (thus invalidating it), we applied it to a number of countries for which herd immunity is not assumed at all: China, France, Germany, United Kingdom, New Zealand and Spain. The reported cases in each these countries has an initial peak. We determined the parameters associated with the best-fitting curve if we fit to *only* that first peak. We used the same optimisation approach as described earlier. For these countries, we enforced a minimum value for $t_g$ of 3 days. In a Reed-Frost model assuming constant $R_{0\_e}$, the ratio of the incline over the decline is never less than 1 – case numbers go down more rapidly than they go up. The lower the value of $R_{0\_e}$, the smaller this difference. Many comparator countries have a slower rate of decline of cases after the first peak than the rate of increase before the first peak – because the Reed-Frost model with herd immunity does not describe well their first peaks. As we try to apply the Reed-Frost model with constant $R_{0\_e}$ and p, lower values of $R_{0\_e}$ result in better fits as a consequence of this curve shape – hence a lower bound restriction is needed. In our main paper, we discuss the value of the resulting parameters and answer the question whether this approach yields viable results across too broad a range of countries.

### 6. Discussion of methodology assumptions and uncertainties

The main finding that in our 10 LMICs of focus, COVID-19 cases have declined as a result of low susceptibility due to high infection levels and immunity, is a reasonable hypothesis to explain the reported cases, appears robust (and thus is worthy of further consideration); it is difficult to explain the observations in another way. However, the parameters determined from our calculations and least-square fits are subject to uncertainty, which we describe here. This is driven by three aspects: (1) limited accuracy of the reported data and specifically, the potential inaccuracy of the assumption that $R_{0\_e}$ and p are constant, (2) the fact that several phenomena have not been incorporated in the model and (3) the covariance between the effective basic reproduction number ($R_{0\_e}$) and infection cycle time ($t_g$) which impact the shape of the curve in a similar way and have a covariance, making it hard to untangle them.

**Firstly, we assume that $R_{0\_e}$ and p are constant.** We know these parameters will not entirely be constant and thus making that assumption is an approximation. In the section around Figure 13, we argued that the maximum variation in $R_{0\_e}$ assumed prior to the peak (if p is constant) was still acceptable and continued to generate parameter values that do not change our key conclusions.

The deviation can also come from a variation in p. We know that the number of tests done for every positive case found (a proxy for p)[11] has varied over time. However, at very low numbers of tests per positive case, the relationship between this number and the detection rate breaks down: if one doubles the number of tests (and thus – everything else held constant – increases the detection rate), at very low numbers the proportion of tests yielding a positive result may not change in line with the detection rate improvement. With many countries being in this low range, at least for part of the period studied, number of tests per positive case is not a sufficient proxy to model p dynamically. In the absence of a credible alternative assumption to model p dynamically, keeping p constant is a prudent choice: given its impact on the curve stretching it vertically, one can make any dataset fit a particular curve, if one allowed any function of p over time. Keeping p constant avoids having too many free parameters, which might lead to good fits even if the model incorrectly describes the disease dynamics.[12]

The high goodness-of-fit (with few degrees of freedom) is the main measure that indicates our assumptions are reasonable. That said, we did do one additional check. In Figure 14 below, we mapped, on log-lin scale the reported case numbers and the reported deaths in all 10 LMICs and, in Figure 15, the 6 comparison countries. As one can see, for each of the 10 selected LMICs, these two curves look very much like each other – even for countries with low absolute numbers of daily new cases and deaths.[13] If the detection rate for cases, p, the ratio of reported cases to actual cases, were to vary wildly, one might expect that the detection/reporting rate for deaths might also be inconsistent over time, and there would be no reason to believe that the two detection rates would vary in the same way over time. A

---

[11] P and the number of tests per positive case are correlated, but not linearly: whilst the number of tests per positive case has halved/ doubled over the period in which we derive the parameter value, this translates in a much smaller variation in p. See https://www.medrxiv.org/content/10.1101/2020.05.12.20098889v1.full.pdf for insights into this link..

[12] We did run a few tests to confirm that the potential range for p (if that were to explain the slight lack of linearity on log-lin scale) does not substantially impact our parameters and findings and found this impact to be very limited.

[13] This is not necessarily the case for the 6 comparator countries. Notably, the widening of the gap between the 2 curves in the UK, France and Germany, could point towards a more expansive testing approach catching more, less-critical cases.

simpler and more reasonable explanation is that the two detection rates – for cases (p) and for deaths – do not vary wildly, which supports the reasonableness of the approximation that the two rates, including p, are constant over time.

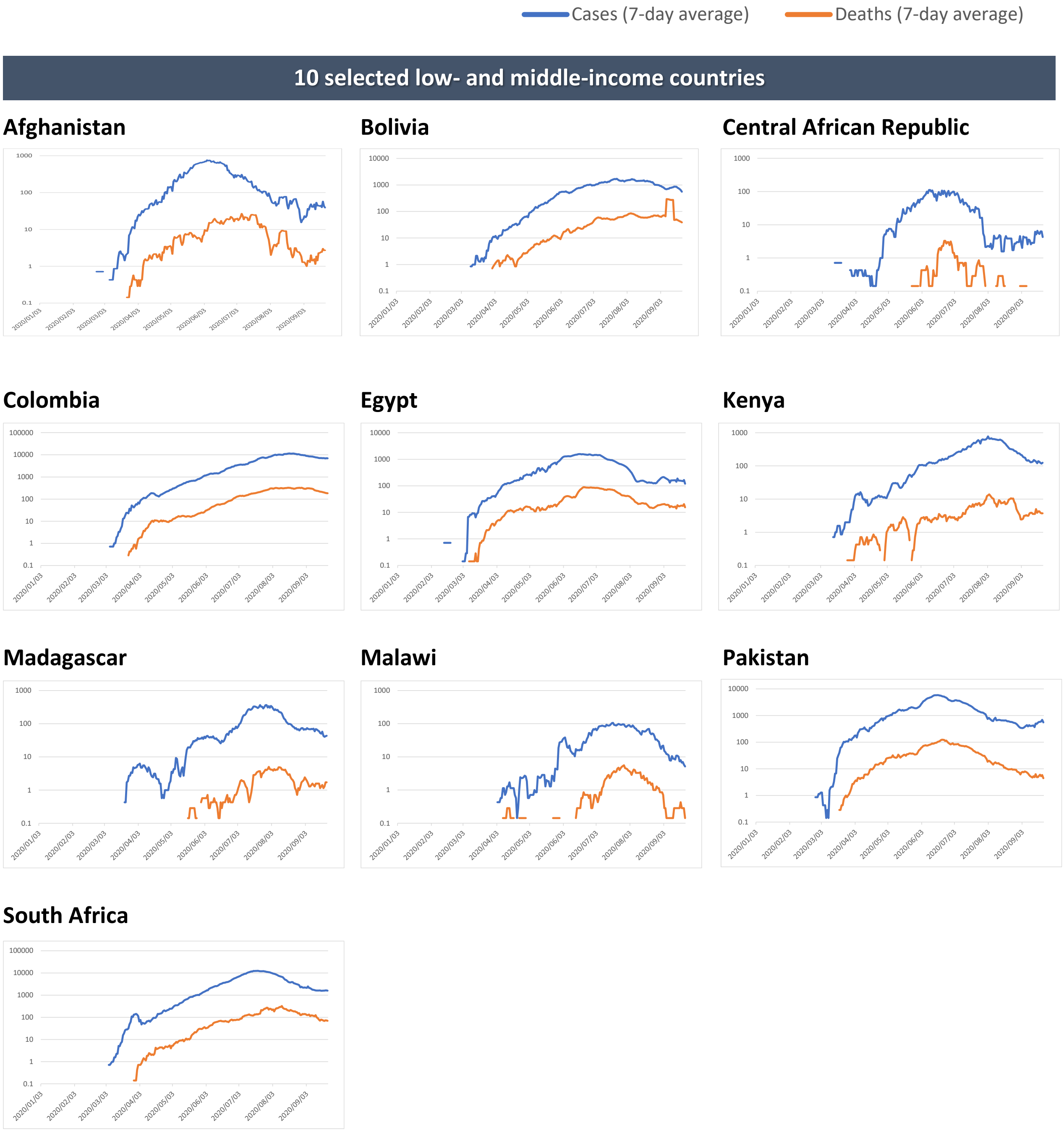

*Figure 14: Reported cases and reported deaths for 10 selected LMICS on log-lin scale*

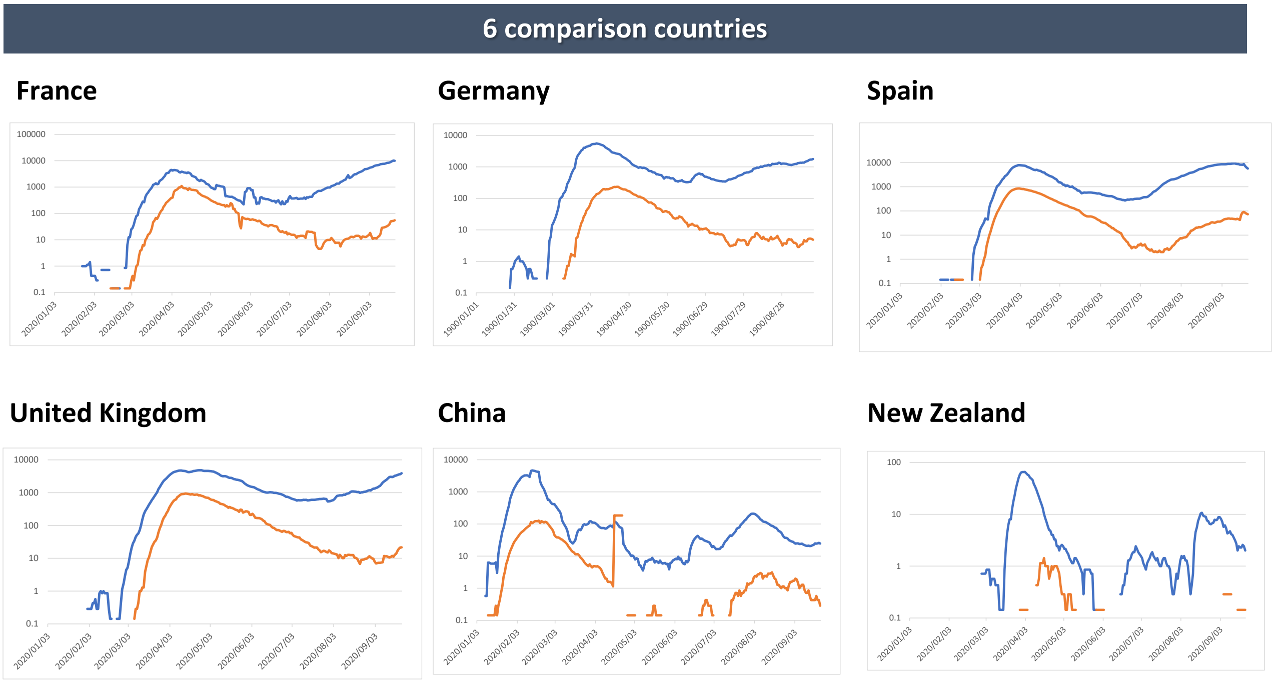

*Figure 15: Reported number of cases and reported number of deaths in 6 comparison countries, on log-lin scale*

**Secondly, as we saw when we described the equations and the effect of different parameters on the curve, $R_{0\_e}$ and $t_g$ are the two parameters that are the hardest to separate.** It is possible to separate them – and our analytical determination for South Africa illustrates this – but it requires good quality data and even then, this approach starts to break down for values for $R_{0\_e}$ below ~ 1.5. However, the fact that their relation uniquely defines the doubling time (derived from the exponential growth rate) means that any reasonable combination yields the exact same pace of the outbreak which reduces the relevance and necessity of exact untangling for understanding the curve writ large. To determine exactly where in the possible range the parameters really sat, one of them needs to be confirmed fully empirically which to date, has not been done.

This means that the values of parameters should be considered order-of-magnitude parameters; especially in situations of a relatively low $R_{0\_e}$ and/ or where poor data quality makes analytical determination of $R_{0\_e}$ and $t_g$ impossible and one needs to rely on regression fitting.

**Thirdly, we will consider heterogeneity.** We are assuming a homogenous society – which is a crude approximation. What follows here, is the mathematical explanation of the inaccuracy that we would introduce in our parameters in three separate cases in which we falsely assumed homogeneity (for two different types of heterogeneity).

Firstly, in a highly interactive society with a few people who have a high infection risk and a lot of people with a relatively low infection risk (a few 'superspreaders' and many, many 'wallflowers'), the people with a high infection risk will get infected and thus immune first – and thus are removed from the pool of 'susceptible' individuals. As a result, the $R_{0\_e}$ of the society will decrease over time, making the exponential growth rate before the peak larger than the exponential decline rate after the peak. This was not observed in South Africa (where we saw assuming a constant $R_{0\_e}$ is a valid assumption), but a weak effect in this type of inhomogeneity could result in an underestimation of $R_{0\_e}$.

Another possibility is that there are several groups in society with almost no interaction and a different level of infection due to different abilities or willingness to socially distance. Conceptually, this would be a few 'subsystems' that each experience their outbreak relatively independently of one another. This will result in similar effect; the exponential growth rate before the peak will be larger than the exponential decline rate after the peak. Also, this will result in peak widening. Again, this was not observed in the SA data so it is likely not an important effect. However, it could result in an underestimation of $R_{0\_e}$.

**The best way to solve this issue is by accurately determining one of the outbreak parameters** – that would allow one to 'lock' that parameter in and determine the other parameters in relation to this exact known value. $T_g$ could be determined from detailed and diligent tracking and tracing data. Alternatively, the total number of infected people could be determined from testing for immunity (serological detection of antibodies in blood; or T cell immunity testing if one wants to include people with a cross-reactive T-cell immunity response – which would most likely confer partial immunity). Of these options, we expect that determining the average $t_g$ will be most accurate.

### 7. Source and approach to determining the derived case fatality rate in Bolivia, Colombia and South Africa

As we discuss in our companion article, we ran a comparison between expected infection fatality rates (IFR), reported fatalities and the infection fatality rate that would be associated with our findings. To do this, we used the following approach:

Firstly, we derived the expected IFR by applying the latest estimates for age-specific IFRs to the breakdown of the population of each of the countries into these age categories. This results in an IFR that adjusts for the country-specific age profile (taking into account the often-cited ‘youthfulness’ of LMICs) and assumes the age-specific IFRs to be constant across countries. For the age-specific IFRs, we used the estimates in Table 2.[14]

*Table 2: Age-specific IFRs*

| **Age category** | **Total, weighted average IFR** |
|---|---:|
| 0-34 | 0.01% |
| 35-44 | 0.06% |
| 45-54 | 0.20% |
| 55-64 | 0.70% |
| 65-74 | 2.20% |
| 75-84 | 7.30% |
| 85+ | 27.10% |

Subsequently, we took the reported deaths from COVID-19 in three countries (Bolivia, Colombia and South Africa – for which information on excess mortality was available), for the period for which excess mortality information was available. We attributed all excess mortality from natural causes to COVID-19, to get an ‘upper band’ estimate of deaths from COVID-19. Lastly, we divided this over the number of infections over that same period, as they result from applying our derived parameters.

[14] A.T. Levin et al. “Assessing the Age Specificity of Infection Fatality Rates for COVID-19: Meta-Analysis & Public Policy Implications” - medRxiv 2020.07.23.20160895 doi: https://doi.org/10.1101/2020.07.23.20160895.

### 8. Centre averaging method to approximate the number of immune people during an infection cycle

The model assumes that the number of infected people is constant during one single time step. As a result, we also assume that the amount of immune people is constant during a time step. Obviously, that is not true, especially when the number of daily infected people is very high. In fact, the way in which the number of infected people during a time step is calculated, can influence the predicted outbreak dynamics. We solved this problem by calculating a low estimate and a high estimate of the of the amount people in the next time step. For the high estimate of $n_{i+1}$ (which we call $n_{i+1}^{+}$), we assume that the amount of immune people during a time step is the amount of immune people at the beginning of that time step:

$$n_{i+1}^{+} = n_i R \left(1 - \frac{n_{immune,i}}{n_{tot}}\right)$$

where $n_{immune,i}$ is the number of immune people at the moment i. Calculating the low estimate of $n_{i+1}$ (which we call $n_{i+1}^{-}$), is slightly less straightforward: for this we need to know the number of immune people at the end of a time step. We cannot calculate this however, since we do not yet know how many people will be infected during a time step. However, since we are dealing with a linear equation in $n_{i+1}$, this can be solved analytically:

$$n_{i+1}^{-} = n_i R \left(1 - \frac{n_{immune,i+1}}{n_{tot}}\right)$$

$$n_{i+1}^{-} = n_i R \left(1 - \frac{n_{immune,i} + t_c n_{i+1}^{-}}{n_{tot}}\right)$$

$$n_{i+1}^{-} = n_i R \left(1 - \frac{n_{immune,i}}{n_{tot}}\right) - n_i R \frac{t_c n_{i+1}^{-}}{n_{tot}}$$

$$n_{i+1}^{-}(1 + \frac{t_c n_i R}{n_{tot}}) = n_i R \left(1 - \frac{n_{immune,i}}{n_{tot}}\right)$$

$$n_{i+1}^{-} = \frac{n_i R \left(1 - \frac{n_{immune,i}}{n_{tot}}\right)}{1 + \frac{t_c n_i R}{n_{tot}}}$$

Now we assume that the best estimate for the number of infected people at the end of step i+1 is the average of $n_{i+1}^{-}$ and $n_{i+1}^{+}$:

$$n_{i+1} = (n_{i+1}^{-} + n_{i+1}^{+})/2$$

We compared the results using the forward (low estimate), backward (high estimate) and centre averaging method and found only small variations. This indicates that whilst centre average (as used) is still the most accurate approximation, the results are not very sensitive to this as opposed to using forward or backward averaging.

### 9. Derivation of the equation for the relationship between $R_{0\_e}$ and $t_g$

Long before the peak, there is exponential growth. The general equation for exponential growth is:

Eq 1) $n = a.e^{\frac{t}{\tau}}$

At t1 there are n1 cases and at t2 there are n2 cases. Substituting in equation 1 gives:

$$n_1 = a.e^{\frac{t_1}{\tau}}$$

$$n_2 = a.e^{\frac{t_2}{\tau}}$$

Choose time step t2 in such a way that t2=t1+ Δt, where Δt = $t_g$.

From the outbreak equation, the ratio of n1 and n2 is given:

$$n_2 = R_{0\_e}.n_1\left[1 - \frac{\int n\,dt}{N_{tot}}\right] = R_{0_e}.n_1$$

$$R_{0\_e} = \frac{n_2}{n_1} = \frac{e^{\frac{t_2}{\tau}}}{e^{\frac{t_1}{\tau}}} = e^{\frac{t_2 - t_1}{\tau}} = e^{\frac{t_g}{\tau}}$$

From which follows Eq 2) $\frac{t_g}{\tau} = \ln R_{0\_e}$

Now choose t3 in such a way that between t1 and t3, the number of cases double:

$$n_1 = a.e^{\frac{t_1}{\tau}}$$

$$n_3 = a.e^{\frac{t_3}{\tau}}$$

We define doubling time $t_d$ where $t_d = t_3 - t_1$ and $\frac{n_3}{n_1} = 2$

Eq 3) $\frac{t_d}{\tau} = \ln 2$

From (eq 2) and (eq 3):

$$\tau = \frac{t_g}{\ln R_{0\_e}} = \frac{t_d}{\ln 2}$$

$$t_d = \frac{\ln 2}{\ln R_{0\_e}} t_g$$
